# Monitoring Report: Respiratory Viruses - February 2026 Data

**DOI:** 10.1101/2024.11.21.24317745

**Authors:** Samuel Gratzl, Karen Gilbert Farrar, Duy Do, Nina Masters, Brianna M Goodwin Cartwright

**Author notes:** Denotes co-first author (Samuel Gratzl, Karen Gilbert Farrar). Corresponding author: Brianna M Goodwin Cartwright Truveta, Inc, 1745 114th Ave SE, Bellevue, WA 98004.

## Abstract

**Background:** Few sources regularly monitor hospitalizations associated with respiratory viruses. This study provides current hospitalization trends associated with six common respiratory viruses: COVID-19, influenza, human metapneumovirus (HMPV), parainfluenza virus, respiratory syncytial virus (RSV), and rhinovirus.

**Objective:** This study aims to supplement the surveillance data provided by the CDC by describing latest trends (through March 01, 2026) overall and for each respiratory virus. This study also provides valuable insight into two at-risk populations: infants and children (age 0-4) and older adults (age 65 and over).

**Methods:** Using a subset of real-world electronic health record (EHR) data from Truveta, a growing collective of health systems that provides more than 18% of all daily clinical care in the US, we identified people who were hospitalized between October 01, 2020 and March 01, 2026. We identified people who tested positive for any of the six respiratory viruses within 14 days of the hospitalization. We report weekly trends in the rate of hospitalizations associated with each virus per all hospitalizations for the overall population and the two high-risk sub populations: infants and children and older adults.

**Results:** We included 1,029,468 hospitalizations of 904,312 unique patients who tested positive for a respiratory virus between October 01, 2020 and March 01, 2026.

Overall, respiratory virus-associated hospitalizations declined slightly throughout February 2026. By the last week of the month, respiratory virus hospitalizations accounted for 3.6% of all hospitalizations, down from 4.3% at the end of January (-15.3%). This decline was primarily driven by influenza and COVID (-34.2% and -16.0%), while RSV, rhinovirus, HMPV-, and parainfluenza-associated hospitalizations all remained relatively low and stable across the month.

Among children aged 0-4 years, respiratory virus-associated hospitalizations increased slightly across February, and accounted for 4.3% of hospitalizations by the end of the month (+11.8%). This increase was primarily driven by HMPV, with HMPV-associated hospitalizations more than doubling throughout February (+111.7%), while RSV-, rhinovirus-, influenza-, COVID- and parainfluenza-associated hospitalizations remained stable (+7.6%, +5.4%, 7.5%, +17.9%, +91.6%). Among adults aged 65 years and older, respiratory virus–associated hospitalizations declined modestly, decreasing from 5.3% to 4.4% of all hospitalizations (−17.0%). This decline was primarily driven by reductions in influenza- and COVID-associated hospitalizations, while RSV-associated hospitalizations increased slightly, and other viruses remained low and stable.

Discussion: Respiratory virus–associated hospitalizations declined slightly during February 2026, reflecting a seasonal decrease in respiratory virus activity. Overall, influenza remained the largest contributor to respiratory virus–associated hospitalizations despite declining hospitalization rates. Among children aged 0–4 years, hospitalization rates remained increased slightly, with RSV continuing to represent the largest share of respiratory virus–associated hospitalizations and HMPV-associated hospitalizations more than doubling over the last month. In contrast, older adults experienced modest declines in respiratory virus–associated hospitalizations, driven primarily by decreases in influenza- and COVID-associated hospitalizations. Together, these findings suggest a gradual seasonal decline in respiratory virus burden across the overall population—particularly among older adults. However, RSV, rhinovirus, influenza, and HMPV continue to contribute to hospitalizations in the pediatric population.

**Trends in surveillance:** *Overall population:* 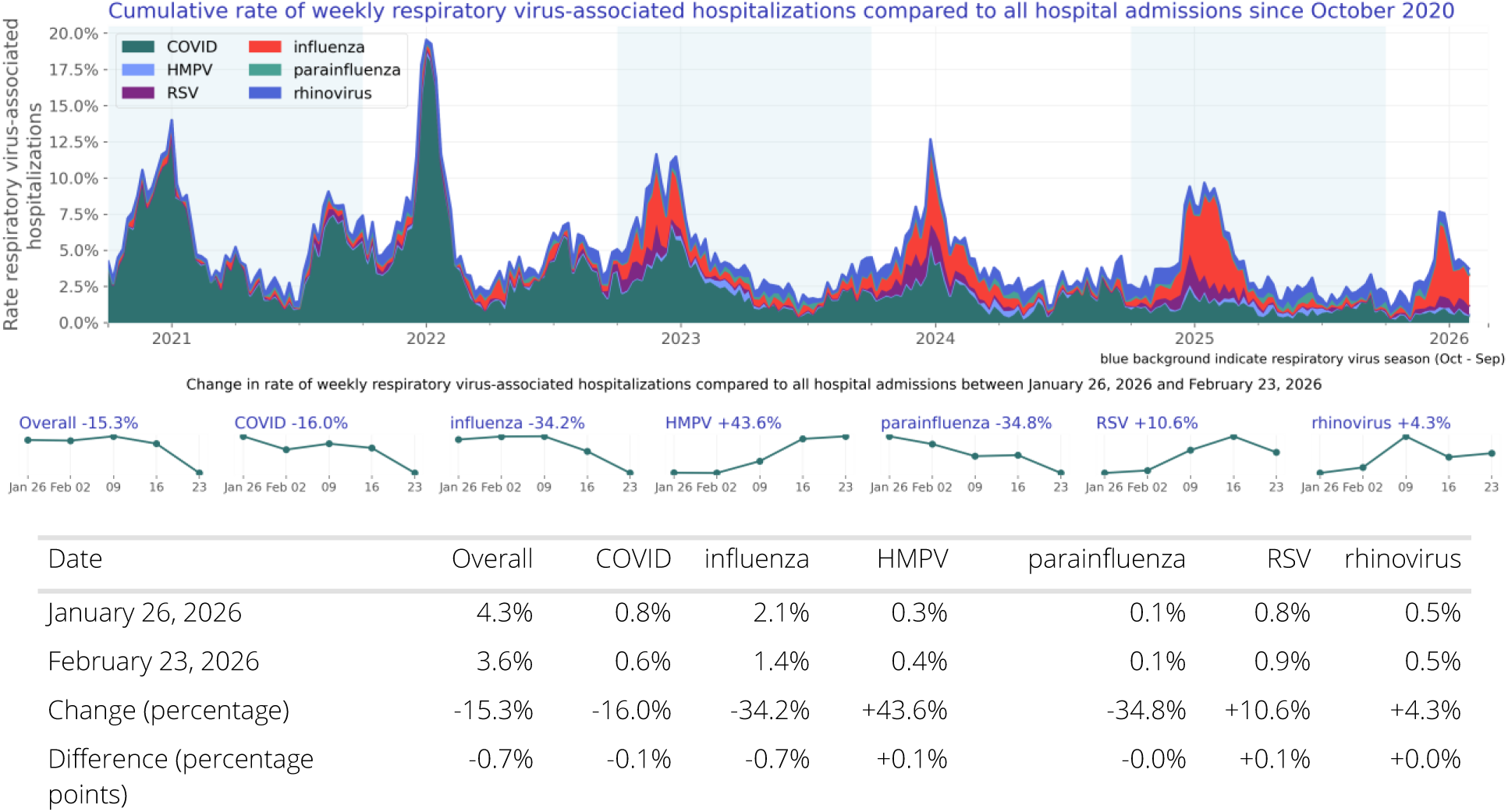

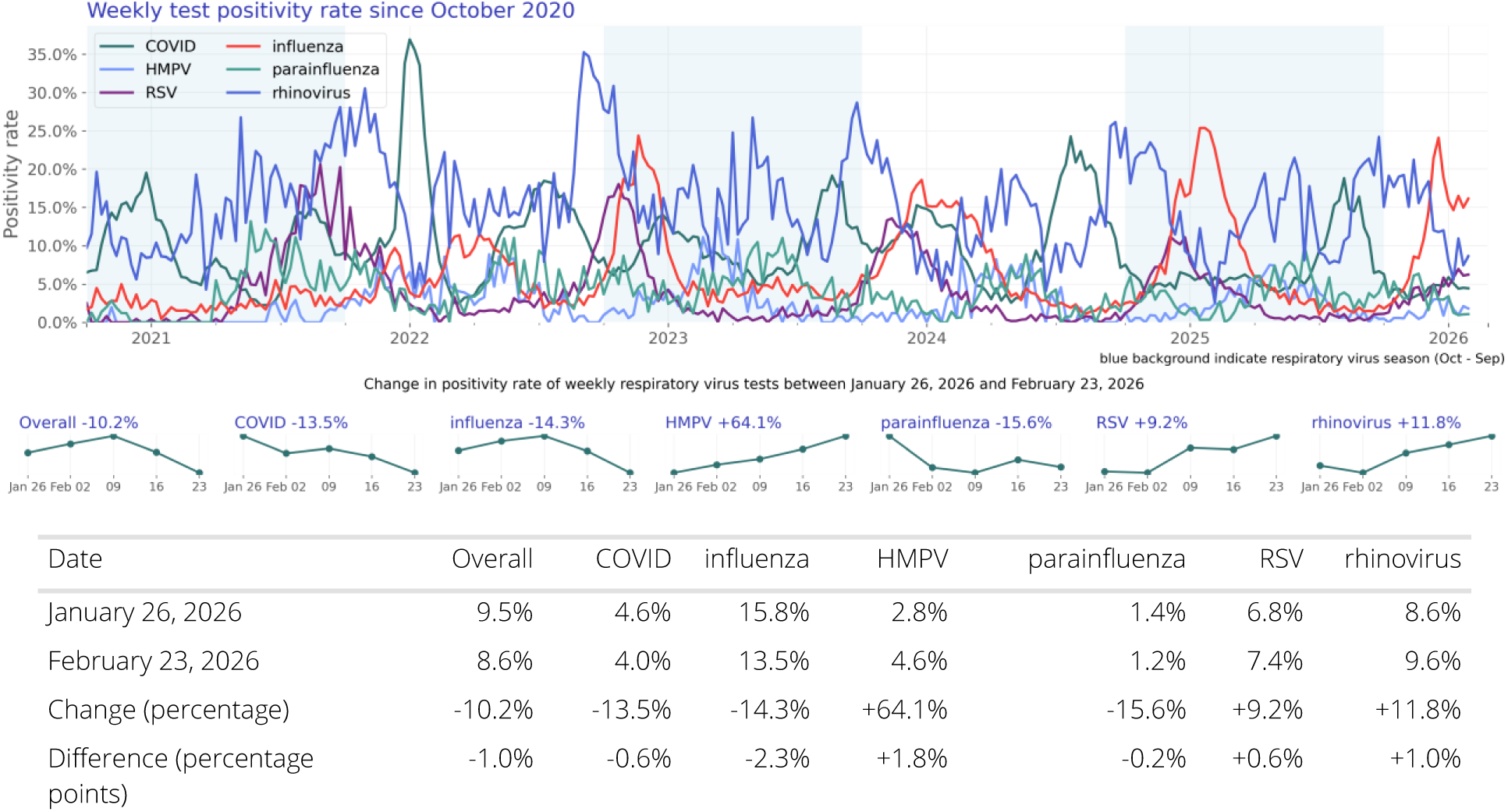 At the end of February 2026, respiratory virus-associated hospitalizations accounted for 3.6% of all hospitalizations, a slight decrease from the end of January 2026 (4.3%, - 15.3%). Influenza-associated hospitalizations decreased from 2.1% to 1.4% of all hospitalizations (-34.2%) and COVID-associated hospitalizations decreased slightly from 0.8% to 0.6% of all hospitalizations (-16.0%). RSV, rhinovirus, HMPV-, and parainfluenza-associated hospitalizations all remained relatively low and stable across the month. Influenza, COVID, and parainfluenza test positivity decreased slightly (-14.3%, -13.5%, - 15.6%, respectively), while rhinovirus, RSV, and HMPV test positivity increased slightly (+11.8%, +9.2%, +64.1%, respectively). The overall findings suggest a slight seasonal decline. Although influenza-associated hospitalizations declined, influenza still accounted for the largest proportion of respiratory virus–associated hospitalizations.

*Infants and children (age 0-4):* 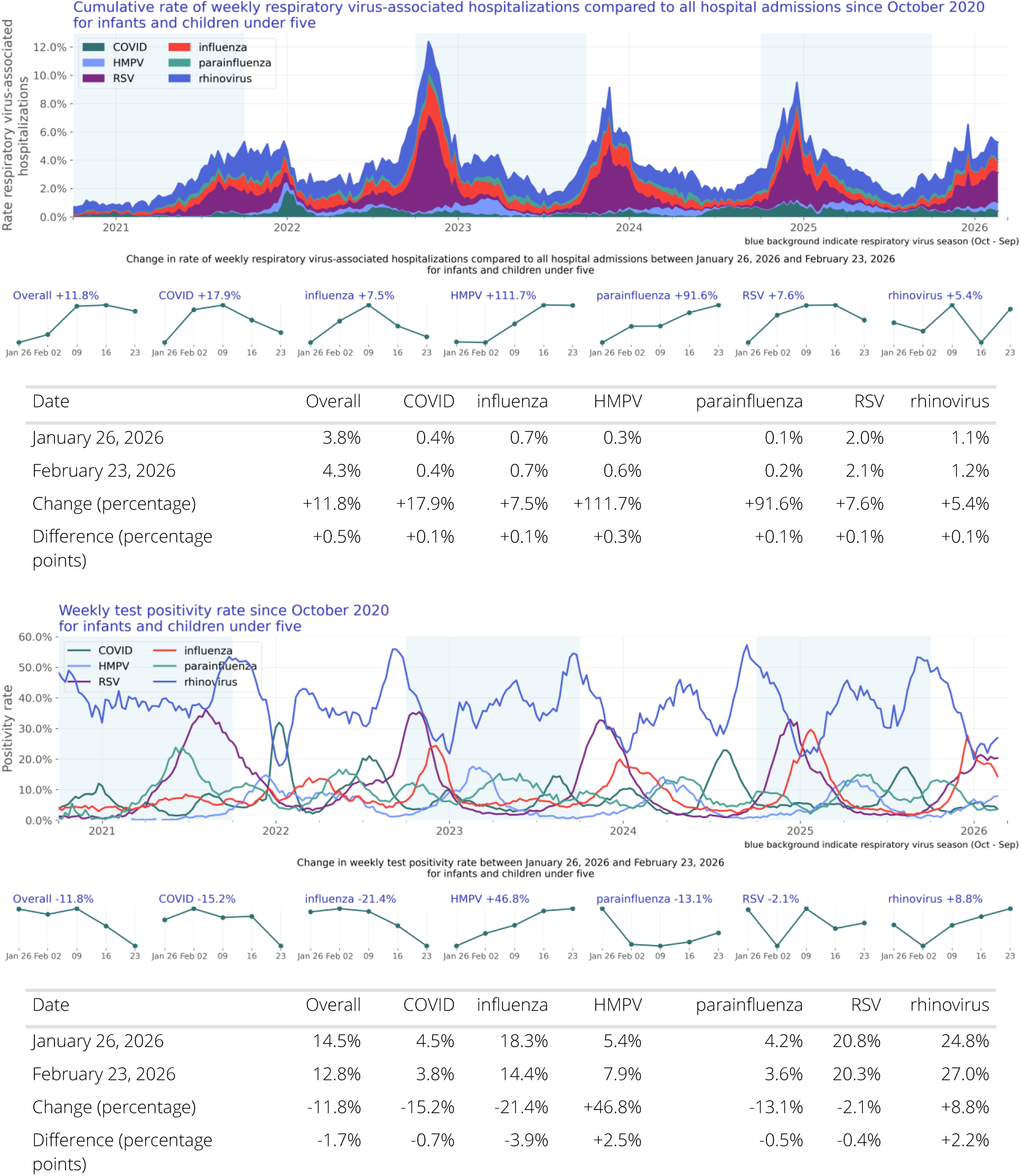 In the population aged 0–4 years, respiratory virus-associated hospitalizations increased slightly throughout February and are now at a rate of 4.3% of all hospitalizations in this age group (+11.8%). This increase was primarily driven by HMPV, with HMPV-associated hospitalizations more than doubling throughout February (+111.7%). RSV-, rhinovirus-, influenza-, COVID- and parainfluenza-associated hospitalizations remained stable (+7.6%, +5.4%, 7.5%, +17.9%, +91.6%). RSV-associated hospitalizations accounted for 2.1% of all hospitalizations by the end of the month, representing the largest share of respiratory-virus associated hospitalizations in this age group. In this pediatric population, rhinovirus and RSV test positivity remained stable (+8.8% and -2.1%, respectively), influenza, COIVD, and parainfluenza test positivity decreased slightly (-21.4%, -15.2%, -13.1%, respectively), and HMPV test positivity increased (+46.8%). Together, these patterns suggest that HMPV drove the increase in respiratory virus activity among children aged 0–4 years, while activity for other viruses remained relatively stable.

*Older adults (age 65 and over):* 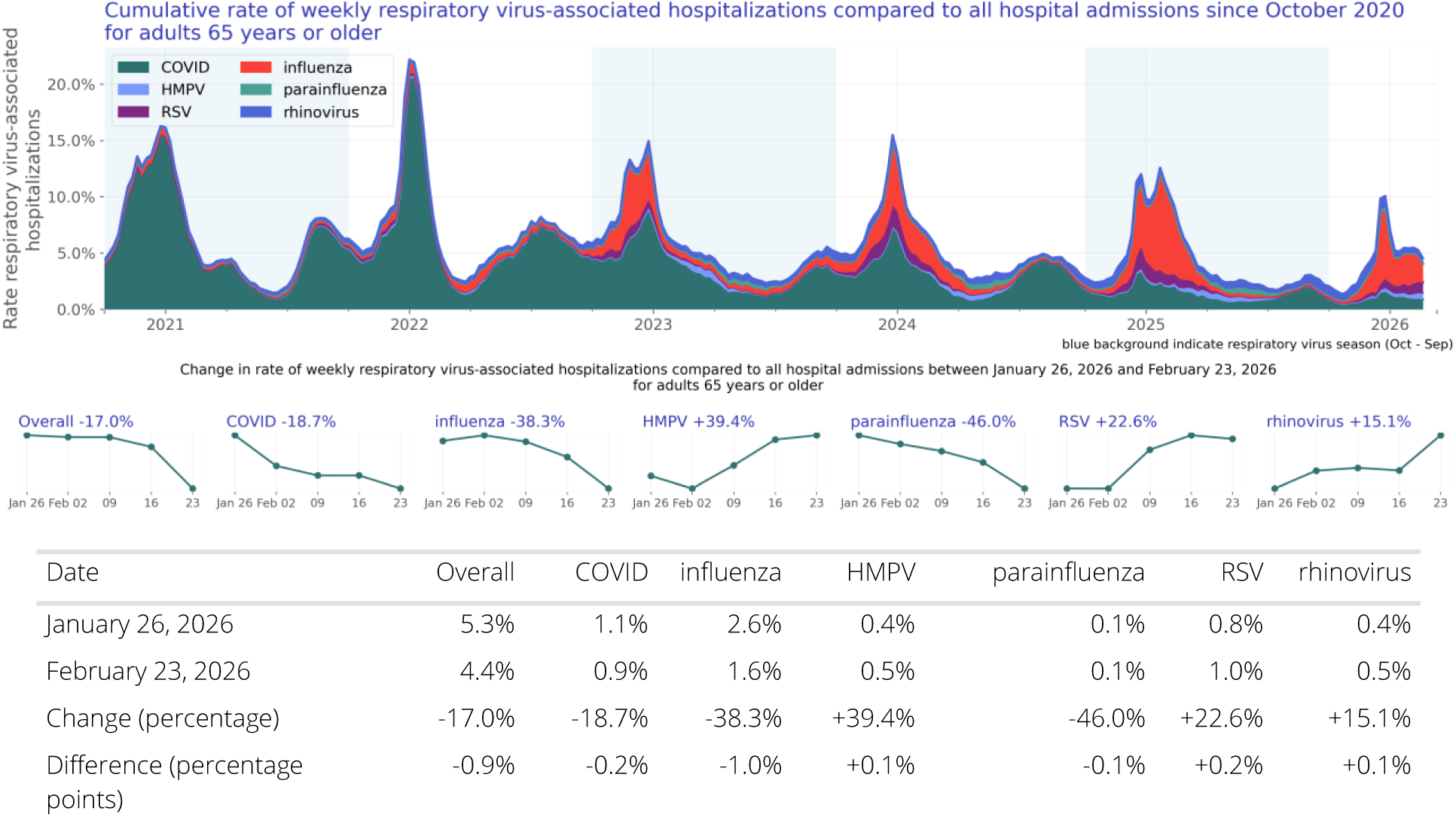

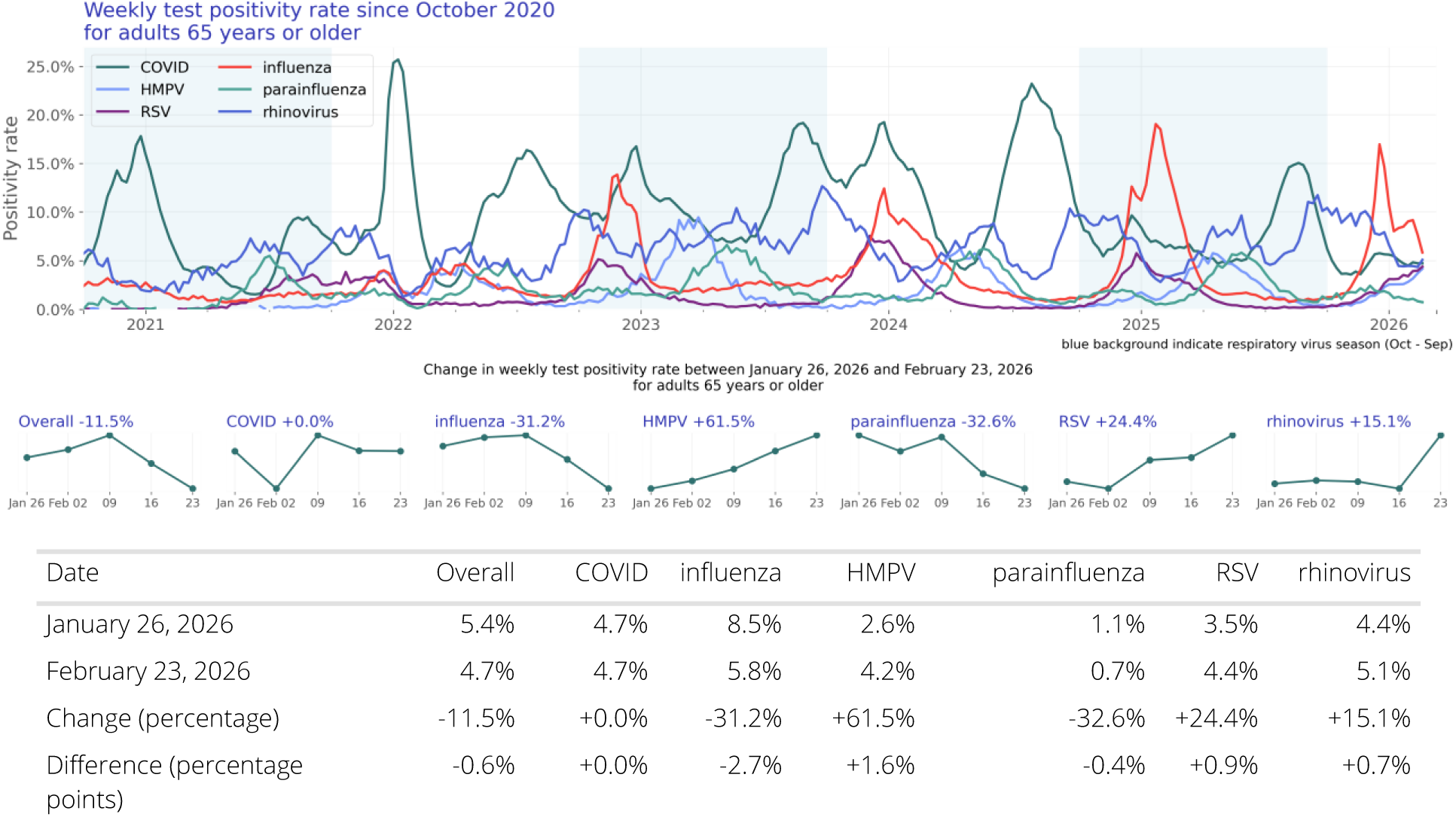 Among adults aged 65 years and older, respiratory virus-associated hospitalizations accounted for 4.4% of all hospitalizations, a slight decrease from the end of January 2026 (5.3%, -17.0%). Influenza- and COVID-associated hospitalizations declined in February (-38.3% and -18.7%, respectively), while RSV-associated hospitalizations increased (+22.6%). HMPV, rhinovirus, and parainfluenza remained low and stable across the month. In this older adult population, influenza and parainfluenza test positivity decreased (-31.2% and -32.6%, respectively), while rhinovirus, RSV, and HMPV test positivity increased (+15.1%, +24.4%, +61.5%). COVID test positivity remained stable across the month (+0.0%). Overall, these findings indicate a slight decrease in respiratory virus-associated hospitalizations among older adults in February, driven primarily by declines in influenza- and COVID-associated hospitalizations.

## Introduction

COVID, influenza, and RSV account for a large proportion of hospitalizations related to respiratory illnesses in the United States. To provide a more complete understanding of hospitalizations related to respiratory viruses, we have also included other viruses known to cause respiratory illness such as human metapneumovirus (HMPV), parainfluenza, and rhinovirus. Each of these viruses can lead to hospitalization and death especially in vulnerable populations, such as infants, children, and older adults (Pastula et al., 2017; Shi et al., 2017, Centers for Disease Control and Prevention 2023a, Smits et al., 2023). Representative and timely data to proactively monitor infections are scarce.

Respiratory viruses are also a major source of infection and hospitalizations in older adults (defined here as patients 65 years of age or older). Incidence has been estimated between 3-10% annually for RSV in older adults (Boyce et al., 2000) and 8-10% for influenza in adults (Tokars et al., 2018). Often, as is the case with influenza, older adults are at higher risk for hospitalization and death than other age groups (Czaja et al. 2019). There are comorbidities that are associated with increased hospitalization risk for older adults, such as congestive heart failure and chronic lung disease (Lee et al., 2013). Further, asthma, COPD, and congestive heart failure can exacerbate respiratory virus infections. Here we report counts for a selection of high-risk medical conditions such as chronic lung diseases and asthma.

Incidence rates in infants and children (defined here as individuals less than five years of age) with respiratory virus infections are higher than other age groups, except adults 65 and older (Centers for Disease Control and Prevention, 2023c; Centers for Disease Control and Prevention, 2023d). In the future, we plan to include high-risk comorbid states, such as congenital heart disease, preterm birth, and cystic fibrosis (Committee on Infectious Diseases and Bronchiolitis Guidelines Committee et al., 2014).

It is important for public health experts and clinical providers to understand the trends in these infections to inform decisions about public health, clinical care, and public policy. Connecting population-level trends with granular clinical information available in Truveta Studio can be very useful to more deeply understand which cohorts are most impacted.

This report is intended to supplement the surveillance data provided by the CDC (Centers for Disease Control and Prevention, 2023b). This report includes additional independent data and clinical detail that is not captured in other reports.

## Methods

### Data

We evaluated respiratory virus-associated hospitalization trends of six common respiratory viruses: COVID-19, influenza, human metapneumovirus (HMPV), parainfluenza virus, respiratory syncytial virus (RSV), and rhinovirus between October 01, 2020 and March 01, 2026 using a subset of Truveta Data. Truveta provides access to continuously updated, linked, and de-identified electronic health record (EHR) from a collective of US health care systems that provide 18% of all daily clinical care in the US, including structured information on demographics, encounters, diagnoses, vital signs (e.g., weight, BMI, blood pressure), medication requests (prescriptions), medication administration, laboratory and diagnostic tests and results (e.g., COVID tests and values), and procedures. Updated EHR data are provided daily to Truveta by constituent health care systems. The data used in this study was provided on March 11, 2026 and included de-identified patient care data primarily located across ten states: California, Florida, Texas, New York, Illinois, Virginia, Washington, Michigan, Ohio, and Arizona.

### Population

We identified hospitalized patients who tested positive for one of the selected respiratory viruses within 14 days before or during the hospitalization. Positive lab results were identified using LOINC codes. Every respiratory virus-associated hospitalization has been grouped such that every hospitalization within 90 days is considered to be the same infection and thus only counted once. To align with seasonality in respiratory transmission, time periods include October 1st through September 30th of the following year.

### Hospitalization rate analysis

Characteristics of respiratory virus-associated hospitalized patients were summarized, including demographics and comorbidities by respiratory virus season. Characteristics are provided for the overall population, individual viruses, and two at-risk sub populations: infants and children (age 0-4) and older adults (age 65 and over).

Respiratory virus-associated hospitalizations rates were summarized over time by virus. The hospitalization rate was calculated weekly as the number of patients with a respiratory virus associated hospitalization divided by the number of patients with a hospital admission in that week. Patients were included in this calculation on the first day of their hospitalization. If their stay was greater than one day, they were not counted on subsequent dates.

Given the unadjusted nature of the data, the rates do not account for undertesting and other variability that exists across patient groups, providers, and systems. For further limitations, see the section below.

### Test positivity rate analysis

Lab results with known results were summarized over time by virus. The positivity rate was calculated monthly as the number of positive lab results divided by the total number of lab results with known results in that month.

Given the unadjusted nature of the data, the rates do not account for undertesting and other variability that exists across patient groups, providers, and systems. For further limitations, see the section below.

## Results

### Overall population

Our study population consists of 1,029,468 hospitalizations of 904,312 unique patients (Table 1).

**Table 1:** Patient characteristics by season.

|  | 2020/2021 | 2021/2022 | 2022/2023 | 2023/2024 | 2024/2025 | 2025/2026 | Overall |
| --- | --- | --- | --- | --- | --- | --- | --- |
| N | 208,226<br>(100.00%) | 219,318<br>(100.00%) | 152,579<br>(100.00%) | 146,852<br>(100.00%) | 124,736<br>(100.00%) | 52,601<br>(100.00%) | 904,312<br>(100.00%) |
| Age Group |  |  |  |  |  |  |  |
| < 6 months | 1,792 (0.86%) | 3,133 (1.43%) | 4,021 (2.64%) | 3,177 (2.16%) | 2,478 (1.99%) | 1,025 (1.95%) | 15,626<br>(1.73%) |
| 6 - <12 months | 785 (0.38%) | 1,506 (0.69%) | 1,963 (1.29%) | 1,693 (1.15%) | 1,517 (1.22%) | 680 (1.29%) | 8,144<br>(0.90%) |
| 1 - <2 years | 1,305 (0.63%) | 2,131 (0.97%) | 2,590 (1.70%) | 2,111 (1.44%) | 2,315 (1.86%) | 1,195 (2.27%) | 11,647<br>(1.29%) |
| 2 - 4 years | 1,400 (0.67%) | 3,121 (1.42%) | 3,745 (2.45%) | 2,585 (1.76%) | 2,906 (2.33%) | 1,406 (2.67%) | 15,163<br>(1.68%) |
| 5 - 17 years | 2,439 (1.17%) | 4,035 (1.84%) | 4,272 (2.80%) | 3,572 (2.43%) | 3,815 (3.06%) | 1,823 (3.47%) | 19,956<br>(2.21%) |
| 18 - 49 years | 49,116<br>(23.59%) | 47,406<br>(21.62%) | 21,949<br>(14.39%) | 20,396<br>(13.89%) | 17,939<br>(14.38%) | 7,711<br>(14.66%) | 164,517<br>(18.19%) |
| 50 - 64 years | 53,873<br>(25.87%) | 46,283<br>(21.10%) | 25,153<br>(16.49%) | 24,681<br>(16.81%) | 21,619<br>(17.33%) | 8,214<br>(15.62%) | 179,823<br>(19.89%) |
| 65 - 74 years | 42,696<br>(20.50%) | 44,029<br>(20.08%) | 30,149<br>(19.76%) | 30,146<br>(20.53%) | 25,534<br>(20.47%) | 10,663<br>(20.27%) | 183,217<br>(20.26%) |
| 75 - 84 years | 35,349<br>(16.98%) | 41,738<br>(19.03%) | 34,678<br>(22.73%) | 35,065<br>(23.88%) | 28,563<br>(22.90%) | 11,895<br>(22.61%) | 187,288<br>(20.71%) |
| 85+ years | 19,471<br>(9.35%) | 25,936<br>(11.83%) | 24,059<br>(15.77%) | 23,426<br>(15.95%) | 18,050<br>(14.47%) | 7,989<br>(15.19%) | 118,931<br>(13.15%) |
| Sex |  |  |  |  |  |  |  |
| Female | 101,925<br>(48.95%) | 112,551<br>(51.32%) | 80,106<br>(52.50%) | 77,121<br>(52.52%) | 66,161<br>(53.04%) | 28,322<br>(53.84%) | 466,186<br>(51.55%) |
| Male | 104,646<br>(50.26%) | 105,090<br>(47.92%) | 71,503<br>(46.86%) | 68,841<br>(46.88%) | 57,970<br>(46.47%) | 23,899<br>(45.43%) | 431,949<br>(47.77%) |
| Other | 3 (0.00%) | 2 (0.00%) | 4 (0.00%) | 1 (0.00%) | 1 (0.00%) | 0 | 11<br>(0.00%) |
| Unknown | 1,652 (0.79%) | 1,675 (0.76%) | 966 (0.63%) | 889 (0.61%) | 604 (0.48%) | 380 (0.72%) | 6,166<br>(0.68%) |
| Race |  |  |  |  |  |  |  |
| White | 134,528<br>(64.61%) | 148,259<br>(67.60%) | 102,058<br>(66.89%) | 97,132<br>(66.14%) | 82,885<br>(66.45%) | 34,669<br>(65.91%) | 599,531<br>(66.30%) |
| Black or African American | 31,628<br>(15.19%) | 31,265<br>(14.26%) | 20,884<br>(13.69%) | 21,537<br>(14.67%) | 17,856<br>(14.32%) | 8,131<br>(15.46%) | 131,301<br>(14.52%) |
| Asian | 4,716 (2.26%) | 4,730 (2.16%) | 4,432 (2.90%) | 3,862 (2.63%) | 3,344 (2.68%) | 1,557 (2.96%) | 22,641<br>(2.50%) |
| Native Hawaiian or Other Pacific Islander | 844 (0.41%) | 925 (0.42%) | 663 (0.43%) | 671 (0.46%) | 675 (0.54%) | 316 (0.60%) | 4,094<br>(0.45%) |
| American Indian or Alaska Native | 1,152 (0.55%) | 1,197 (0.55%) | 824 (0.54%) | 733 (0.50%) | 662 (0.53%) | 261 (0.50%) | 4,829<br>(0.53%) |
| Other Race | 13,692<br>(6.58%) | 11,480<br>(5.23%) | 8,978 (5.88%) | 8,372 (5.70%) | 8,047 (6.45%) | 3,104 (5.90%) | 53,673<br>(5.94%) |
| Unknown | 21,666<br>(10.41%) | 21,462<br>(9.79%) | 14,740<br>(9.66%) | 14,545<br>(9.90%) | 11,267<br>(9.03%) | 4,563 (8.67%) | 88,243<br>(9.76%) |
| Ethnicity |  |  |  |  |  |  |  |
| Hispanic or Latino | 24,264<br>(11.65%) | 19,248<br>(8.78%) | 14,054<br>(9.21%) | 14,224<br>(9.69%) | 14,011<br>(11.23%) | 5,757<br>(10.94%) | 91,558<br>(10.12%) |
| Not Hispanic or Latino | 161,393<br>(77.51%) | 179,073<br>(81.65%) | 124,535<br>(81.62%) | 119,629<br>(81.46%) | 101,364<br>(81.26%) | 43,322<br>(82.36%) | 729,316<br>(80.65%) |
| Unknown | 22,569<br>(10.84%) | 20,997<br>(9.57%) | 13,990<br>(9.17%) | 12,999<br>(8.85%) | 9,361 (7.50%) | 3,522 (6.70%) | 83,438<br>(9.23%) |
| Virus |  |  |  |  |  |  |  |
| COVID | 190,101<br>(91.30%) | 186,074<br>(84.84%) | 86,671<br>(56.80%) | 75,419<br>(51.36%) | 37,889<br>(30.38%) | 10,340<br>(19.66%) | 586,494<br>(64.86%) |
| HMPV | 100 (0.05%) | 2,986 (1.36%) | 4,991 (3.27%) | 4,282 (2.92%) | 5,591 (4.48%) | 2,361 (4.49%) | 20,311<br>(2.25%) |
| RSV | 2,339 (1.12%) | 4,955 (2.26%) | 9,513 (6.23%) | 11,910<br>(8.11%) | 12,774<br>(10.24%) | 6,434<br>(12.23%) | 47,925<br>(5.30%) |
| influenza | 7,275 (3.49%) | 13,765<br>(6.28%) | 34,203<br>(22.42%) | 37,301<br>(25.40%) | 45,183<br>(36.22%) | 22,174<br>(42.16%) | 159,901<br>(17.68%) |
| parainfluenza | 1,526 (0.73%) | 2,446 (1.12%) | 4,630 (3.03%) | 4,937 (3.36%) | 5,635 (4.52%) | 1,934 (3.68%) | 21,108<br>(2.33%) |
| rhinovirus | 6,885 (3.31%) | 9,092 (4.15%) | 12,571<br>(8.24%) | 13,003<br>(8.85%) | 17,664<br>(14.16%) | 9,358<br>(17.79%) | 68,573<br>(7.58%) |
| Comorbidity |  |  |  |  |  |  |  |
| Asthma | 18,004<br>(8.65%) | 22,064<br>(10.06%) | 18,978<br>(12.44%) | 20,146<br>(13.72%) | 19,518<br>(15.65%) | 8,604<br>(16.36%) | 107,314<br>(11.87%) |
| Chronic Lung Disease | 12,918<br>(6.20%) | 14,607<br>(6.66%) | 11,750<br>(7.70%) | 12,190<br>(8.30%) | 12,090<br>(9.69%) | 5,125 (9.74%) | 68,680<br>(7.59%) |
| Chronic Obstructive Pulmonary Disease | 23,282<br>(11.18%) | 32,805<br>(14.96%) | 27,930<br>(18.31%) | 29,762<br>(20.27%) | 26,850<br>(21.53%) | 11,169<br>(21.23%) | 151,798<br>(16.79%) |

#### Hospitalization rate over time

The rate of respiratory virus-associated hospitalizations compared to all hospitalizations is shown in Figure 1. Figure 2 shows the same data stacked to represent the combined impact of the viruses.

**Figure 1:**
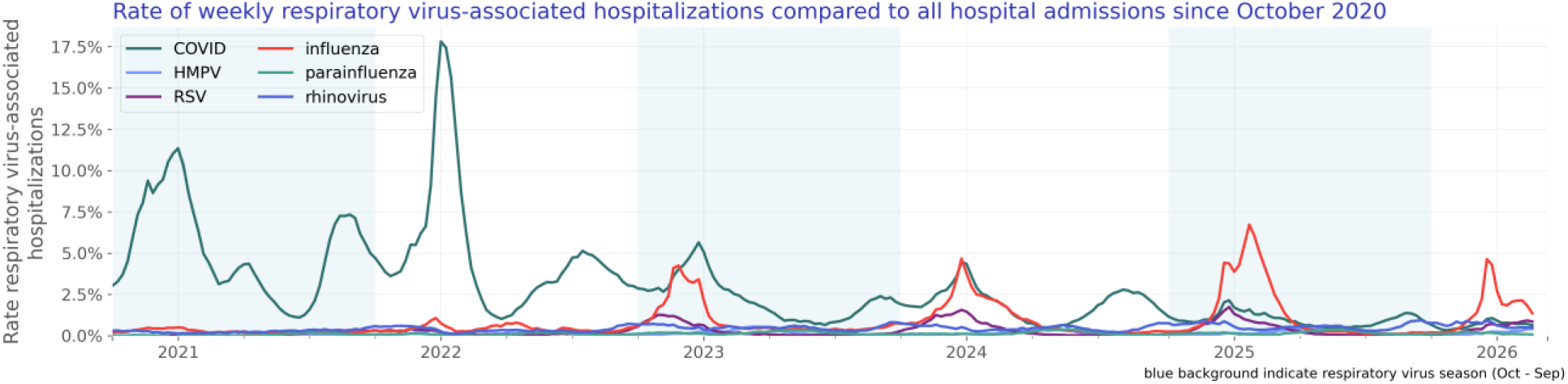
Rate of weekly respiratory virus-associated hospitalizations compared to all hospital admissions since October 2020.

**Figure 2:**
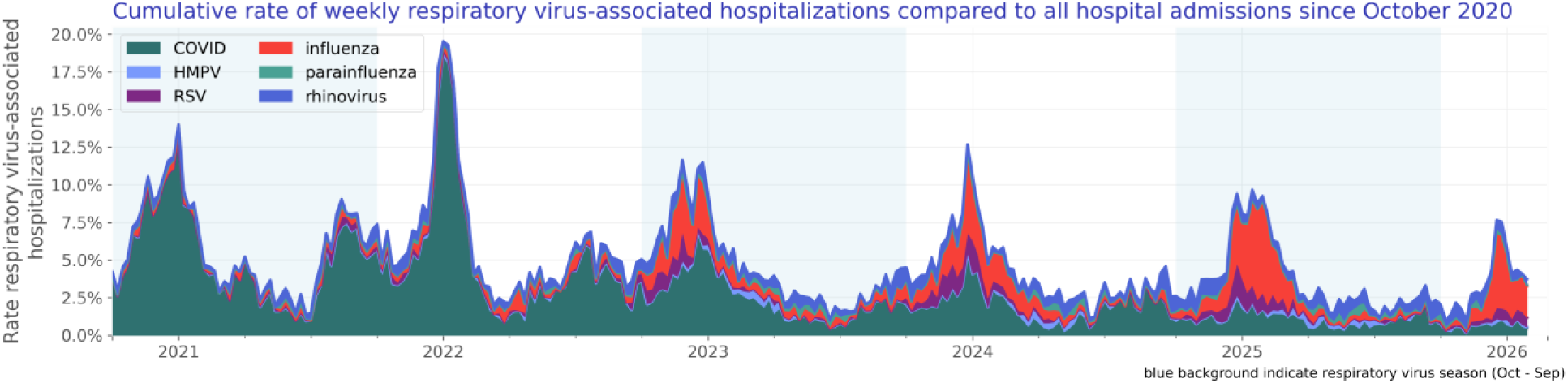
Cumulative rate of weekly respiratory virus-associated hospitalizations compared to all hospital admissions since October 2020.

#### Test positivity rate over time

We included 82,033,479 lab results with known results of which 7,672,331 were positive. The test positivity rate is shown in Figure 3.

**Figure 3:**
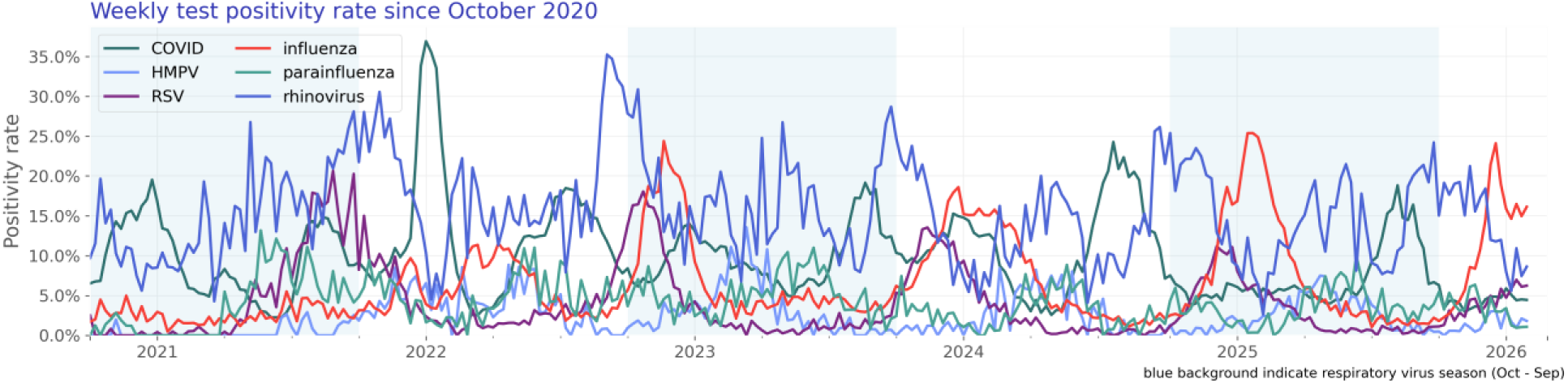
Weekly test positivity rate since October 2020.

### COVID-19

Our COVID study population consists of 610,502 hospitalizations of 586,494 unique patients (Table 2).

**Table 2:** COVID patient characteristics by season.

|  | 2020/2021 | 2021/2022 | 2022/2023 | 2023/2024 | 2024/2025 | 2025/2026 | Overall |
| --- | --- | --- | --- | --- | --- | --- | --- |
| N | 190,101<br>(100.00%) | 186,074<br>(100.00%) | 86,671<br>(100.00%) | 75,419<br>(100.00%) | 37,889<br>(100.00%) | 10,340<br>(100.00%) | 586,494<br>(100.00%) |
| Age Group |  |  |  |  |  |  |  |
| < 6 months | 338 (0.18%) | 865 (0.46%) | 419 (0.48%) | 430 (0.57%) | 386 (1.02%) | 117 (1.13%) | 2,555<br>(0.44%) |
| 6 - <12 months | 60 (0.03%) | 269 (0.14%) | 181 (0.21%) | 218 (0.29%) | 243 (0.64%) | 75 (0.73%) | 1,046<br>(0.18%) |
| 1 - <2 years | 93 (0.05%) | 240 (0.13%) | 135 (0.16%) | 269 (0.36%) | 452 (1.19%) | 139 (1.34%) | 1,328<br>(0.23%) |
| 2 - 4 years | 135 (0.07%) | 349 (0.19%) | 130 (0.15%) | 267 (0.35%) | 615 (1.62%) | 224 (2.17%) | 1,720<br>(0.29%) |
| 5 - 17 years | 1,149 (0.60%) | 1,681 (0.90%) | 569 (0.66%) | 613 (0.81%) | 990 (2.61%) | 309 (2.99%) | 5,311<br>(0.91%) |
| 18 - 49 years | 45,798<br>(24.09%) | 42,262<br>(22.71%) | 11,774<br>(13.58%) | 8,499<br>(11.27%) | 4,340<br>(11.45%) | 1,214<br>(11.74%) | 113,887<br>(19.42%) |
| 50 - 64 years | 50,668<br>(26.65%) | 41,121<br>(22.10%) | 13,726<br>(15.84%) | 11,226<br>(14.88%) | 5,133<br>(13.55%) | 1,378<br>(13.33%) | 123,252<br>(21.02%) |
| 65 - 74 years | 40,179<br>(21.14%) | 39,000<br>(20.96%) | 18,565<br>(21.42%) | 16,358<br>(21.69%) | 7,621<br>(20.11%) | 2,190<br>(21.18%) | 123,913<br>(21.13%) |
| 75 - 84 years | 33,355<br>(17.55%) | 37,236<br>(20.01%) | 23,795<br>(27.45%) | 21,937<br>(29.09%) | 10,729<br>(28.32%) | 2,765<br>(26.74%) | 129,817<br>(22.13%) |
| 85+ years | 18,326<br>(9.64%) | 23,051<br>(12.39%) | 17,377<br>(20.05%) | 15,602<br>(20.69%) | 7,380<br>(19.48%) | 1,929<br>(18.66%) | 83,665<br>(14.27%) |
| Sex |  |  |  |  |  |  |  |
| Female | 92,840<br>(48.84%) | 95,248<br>(51.19%) | 44,863<br>(51.76%) | 38,481<br>(51.02%) | 19,487<br>(51.43%) | 5,371<br>(51.94%) | 296,290<br>(50.52%) |
| Male | 95,764<br>(50.38%) | 89,352<br>(48.02%) | 41,214<br>(47.55%) | 36,451<br>(48.33%) | 18,210<br>(48.06%) | 4,873<br>(47.13%) | 285,864<br>(48.74%) |
| Other | 3 (0.00%) | 2 (0.00%) | 2 (0.00%) | 1 (0.00%) | 0 | 0 | 8 (0.00%) |
| Unknown | 1,494 (0.79%) | 1,472 (0.79%) | 592 (0.68%) | 486 (0.64%) | 192 (0.51%) | 96 (0.93%) | 4,332<br>(0.74%) |
| Race |  |  |  |  |  |  |  |
| White | 124,012<br>(65.23%) | 127,598<br>(68.57%) | 61,426<br>(70.87%) | 52,800<br>(70.01%) | 25,583<br>(67.52%) | 6,948<br>(67.20%) | 398,367<br>(67.92%) |
| Black or African American | 28,477<br>(14.98%) | 26,331<br>(14.15%) | 10,600<br>(12.23%) | 9,441<br>(12.52%) | 4,943<br>(13.05%) | 1,436<br>(13.89%) | 81,228<br>(13.85%) |
| Asian | 4,101 (2.16%) | 3,533 (1.90%) | 2,021 (2.33%) | 1,918 (2.54%) | 1,216 (3.21%) | 343 (3.32%) | 13,132<br>(2.24%) |
| Native Hawaiian or Other Pacific Islander | 769 (0.40%) | 708 (0.38%) | 274 (0.32%) | 259 (0.34%) | 139 (0.37%) | 26 (0.25%) | 2,175<br>(0.37%) |
| American Indian or Alaska Native | 1,034 (0.54%) | 984 (0.53%) | 392 (0.45%) | 358 (0.47%) | 184 (0.49%) | 57 (0.55%) | 3,009 (0.51%) |
| Other Race | 11,669 (6.14%) | 8,415 (4.52%) | 3,639 (4.20%) | 3,305 (4.38%) | 2,281 (6.02%) | 586 (5.67%) | 29,895 (5.10%) |
| Unknown | 20,039 (10.54%) | 18,505 (9.94%) | 8,319 (9.60%) | 7,338 (9.73%) | 3,543 (9.35%) | 944 (9.13%) | 58,688 (10.01%) |
| Ethnicity |  |  |  |  |  |  |  |
| Hispanic or Latino | 22,085 (11.62%) | 15,098 (8.11%) | 6,023 (6.95%) | 5,791 (7.68%) | 3,269 (8.63%) | 764 (7.39%) | 53,030 (9.04%) |
| Not Hispanic or Latino | 147,413 (77.54%) | 153,043 (82.25%) | 72,584 (83.75%) | 62,948 (83.46%) | 31,595 (83.39%) | 8,801 (85.12%) | 476,384 (81.23%) |
| Unknown | 20,603 (10.84%) | 17,933 (9.64%) | 8,064 (9.30%) | 6,680 (8.86%) | 3,025 (7.98%) | 775 (7.50%) | 57,080 (9.73%) |
| Comorbidity |  |  |  |  |  |  |  |
| Asthma | 15,386 (8.09%) | 17,122 (9.20%) | 8,904 (10.27%) | 8,770 (11.63%) | 4,943 (13.05%) | 1,415 (13.68%) | 56,540 (9.64%) |
| Chronic Lung Disease | 11,645 (6.13%) | 12,026 (6.46%) | 6,514 (7.52%) | 6,169 (8.18%) | 3,197 (8.44%) | 964 (9.32%) | 40,515 (6.91%) |
| Chronic Obstructive Pulmonary Disease | 20,590 (10.83%) | 27,186 (14.61%) | 15,892 (18.34%) | 15,478 (20.52%) | 7,582 (20.01%) | 2,136 (20.66%) | 88,864 (15.15%) |

#### Hospitalization rate over time

The rate of COVID-associated hospitalization is shown in Figure 4. Figure 5 shows seasonal trends.

**Figure 4:**
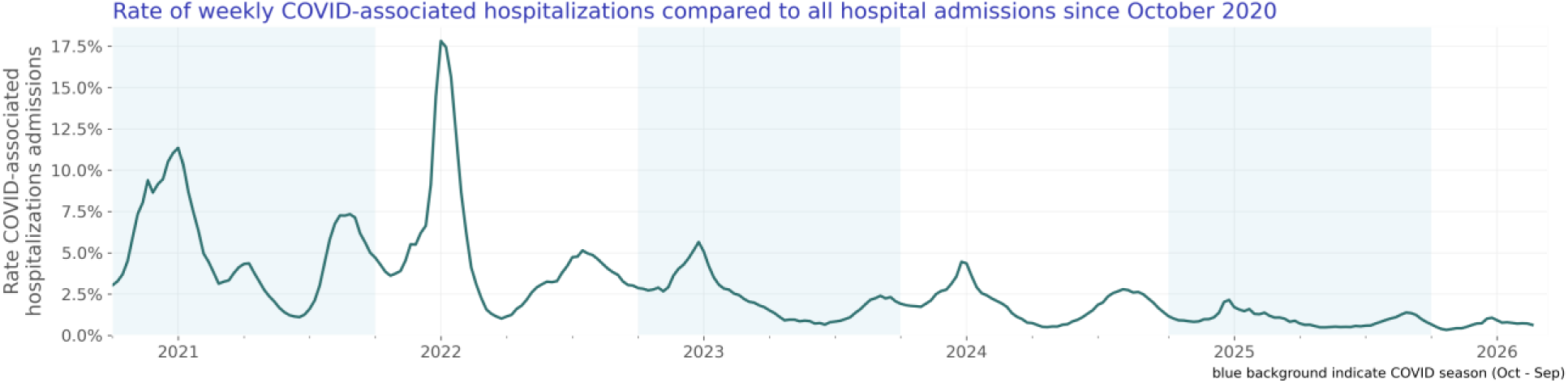
Rate of weekly COVID-associated hospitalizations compared to all hospital admissions since October 2020.

**Figure 5:**
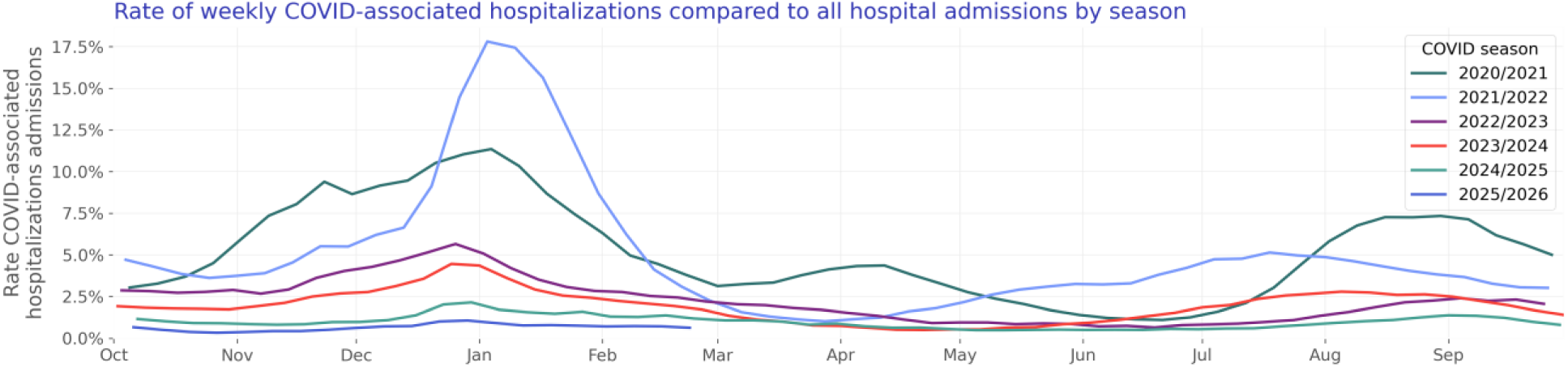
Rate of weekly COVID-associated hospitalizations compared to all hospital admissions by season.

#### Test positivity rate over time

We included 40,309,628 COVID lab results with known results of which 4,341,641 were positive. The COVID test positivity rate is shown in Figure 6. Figure 7 shows yearly trends.

**Figure 6:**
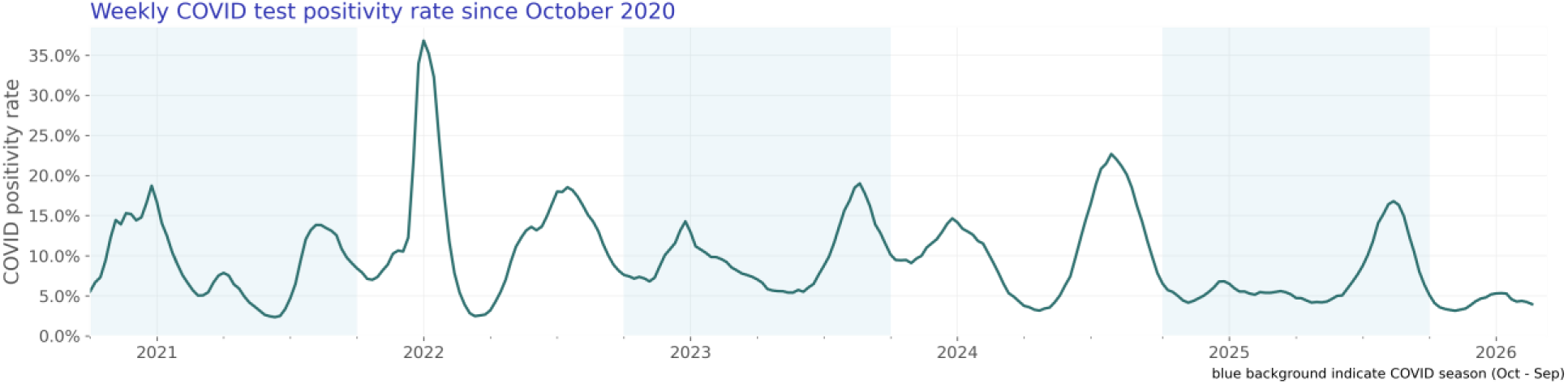
Weekly COVID test positivity rate since October 2020.

**Figure 7:**
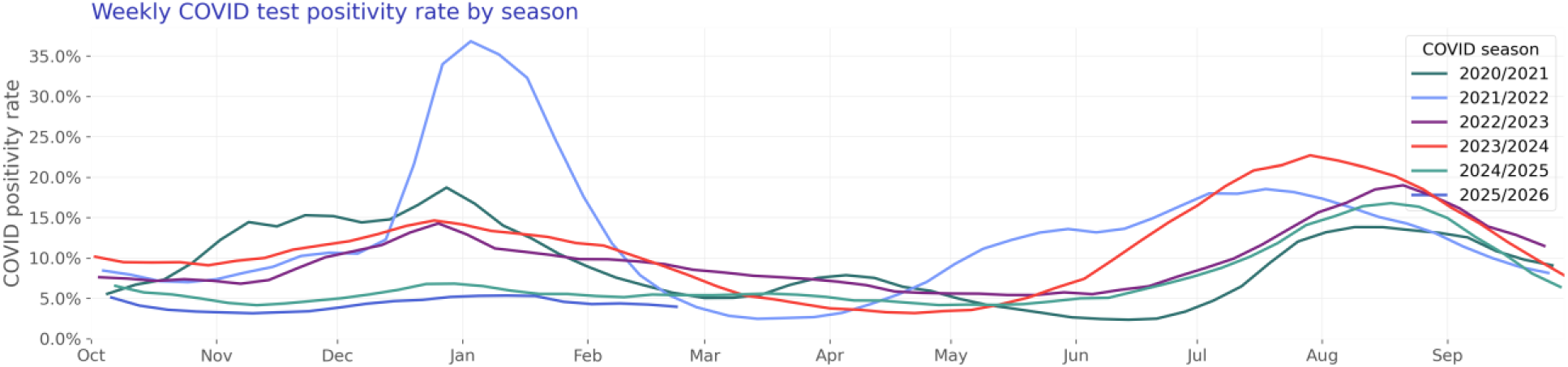
Weekly COVID test positivity rate by season.

### Influenza

Our influenza study population consists of 197,256 hospitalizations of 159,901 unique patients (Table 3).

**Table 3:**
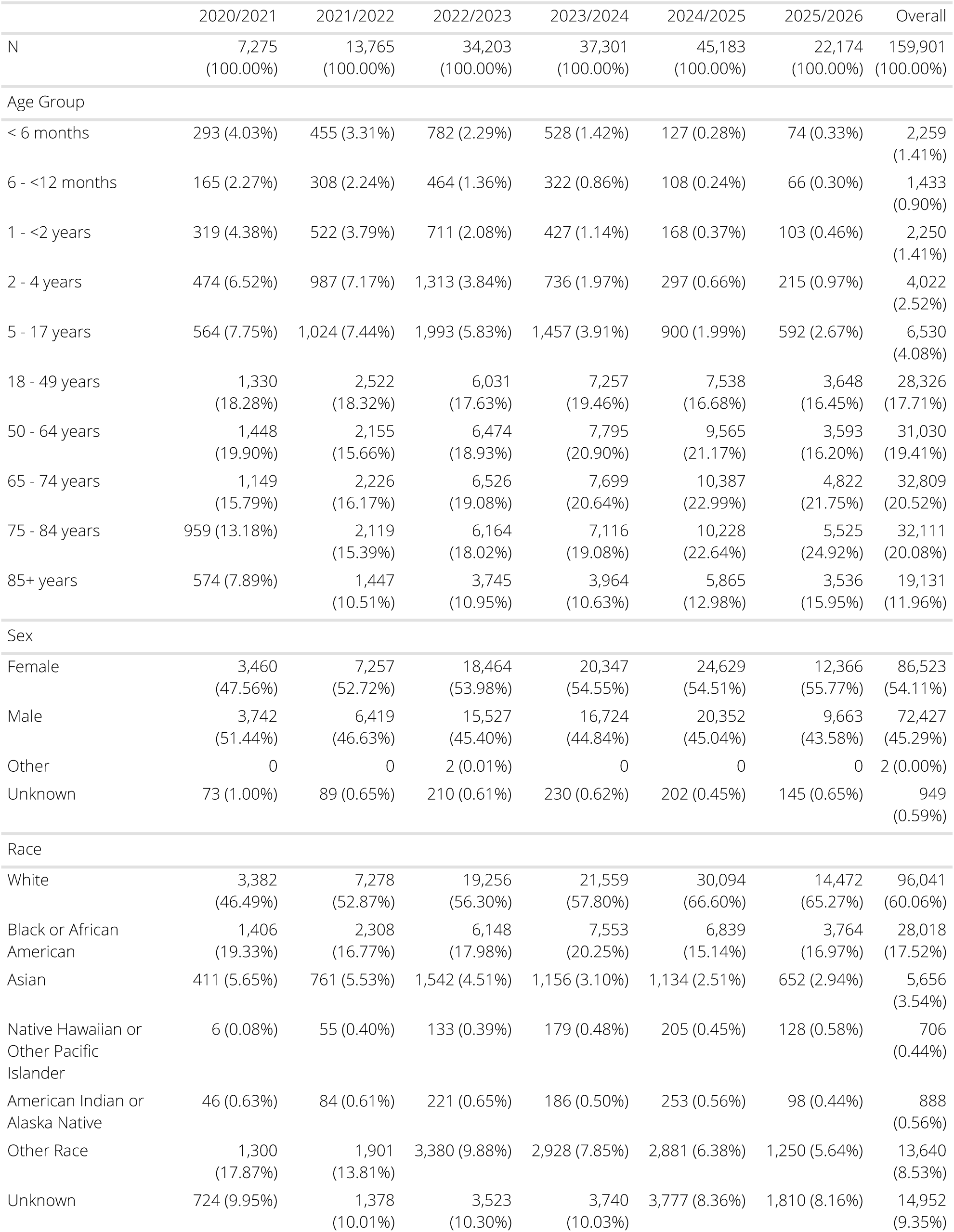
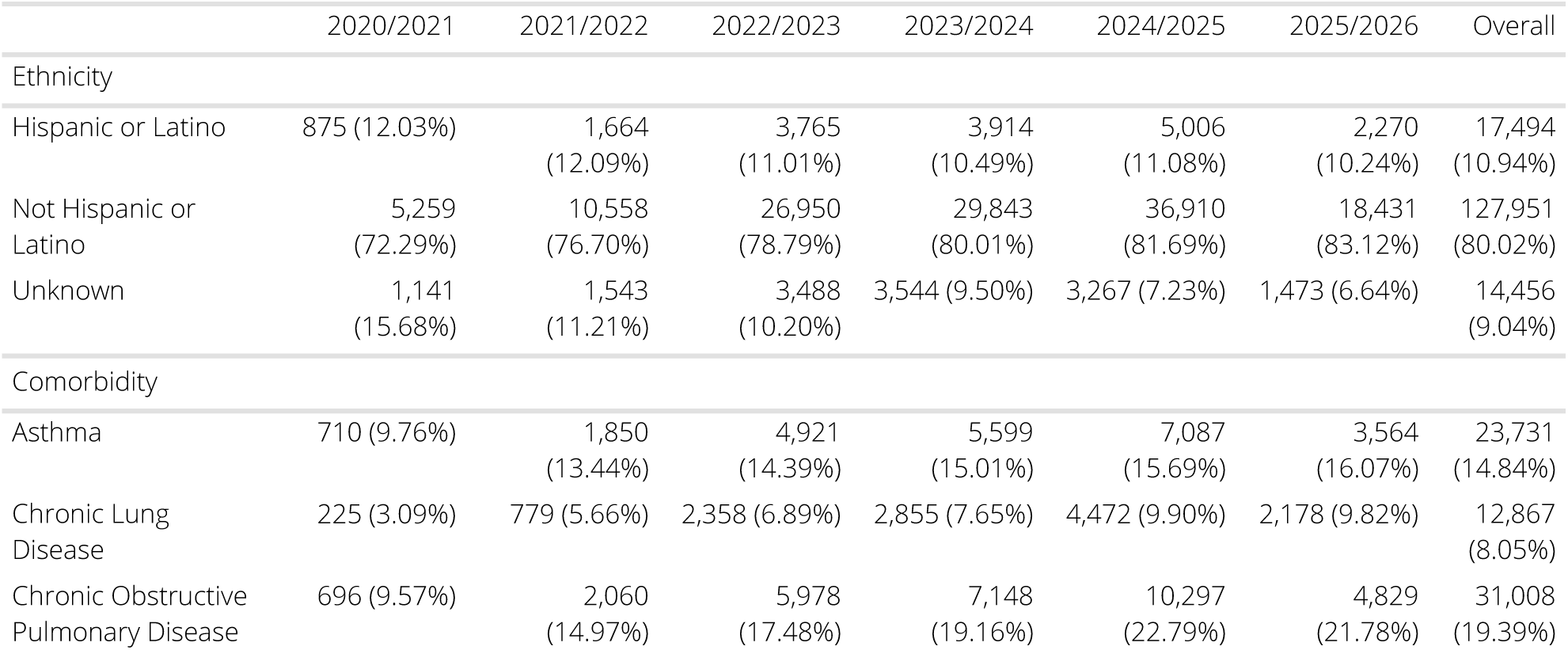
Influenza patient characteristics by season.

|  | 2020/2021 | 2021/2022 | 2022/2023 | 2023/2024 | 2024/2025 | 2025/2026 | Overall |
| --- | --- | --- | --- | --- | --- | --- | --- |
| N | 7,275<br>(100.00%) | 13,765<br>(100.00%) | 34,203<br>(100.00%) | 37,301<br>(100.00%) | 45,183<br>(100.00%) | 22,174<br>(100.00%) | 159,901<br>(100.00%) |
| Age Group |  |  |  |  |  |  |  |
| < 6 months | 293 (4.03%) | 455 (3.31%) | 782 (2.29%) | 528 (1.42%) | 127 (0.28%) | 74 (0.33%) | 2,259<br>(1.41%) |
| 6 - <12 months | 165 (2.27%) | 308 (2.24%) | 464 (1.36%) | 322 (0.86%) | 108 (0.24%) | 66 (0.30%) | 1,433<br>(0.90%) |
| 1 - <2 years | 319 (4.38%) | 522 (3.79%) | 711 (2.08%) | 427 (1.14%) | 168 (0.37%) | 103 (0.46%) | 2,250<br>(1.41%) |
| 2 - 4 years | 474 (6.52%) | 987 (7.17%) | 1,313 (3.84%) | 736 (1.97%) | 297 (0.66%) | 215 (0.97%) | 4,022<br>(2.52%) |
| 5 - 17 years | 564 (7.75%) | 1,024 (7.44%) | 1,993 (5.83%) | 1,457 (3.91%) | 900 (1.99%) | 592 (2.67%) | 6,530<br>(4.08%) |
| 18 - 49 years | 1,330<br>(18.28%) | 2,522<br>(18.32%) | 6,031<br>(17.63%) | 7,257<br>(19.46%) | 7,538<br>(16.68%) | 3,648<br>(16.45%) | 28,326<br>(17.71%) |
| 50 - 64 years | 1,448<br>(19.90%) | 2,155<br>(15.66%) | 6,474<br>(18.93%) | 7,795<br>(20.90%) | 9,565<br>(21.17%) | 3,593<br>(16.20%) | 31,030<br>(19.41%) |
| 65 - 74 years | 1,149<br>(15.79%) | 2,226<br>(16.17%) | 6,526<br>(19.08%) | 7,699<br>(20.64%) | 10,387<br>(22.99%) | 4,822<br>(21.75%) | 32,809<br>(20.52%) |
| 75 - 84 years | 959 (13.18%) | 2,119<br>(15.39%) | 6,164<br>(18.02%) | 7,116<br>(19.08%) | 10,228<br>(22.64%) | 5,525<br>(24.92%) | 32,111<br>(20.08%) |
| 85+ years | 574 (7.89%) | 1,447<br>(10.51%) | 3,745<br>(10.95%) | 3,964<br>(10.63%) | 5,865<br>(12.98%) | 3,536<br>(15.95%) | 19,131<br>(11.96%) |
| Sex |  |  |  |  |  |  |  |
| Female | 3,460<br>(47.56%) | 7,257<br>(52.72%) | 18,464<br>(53.98%) | 20,347<br>(54.55%) | 24,629<br>(54.51%) | 12,366<br>(55.77%) | 86,523<br>(54.11%) |
| Male | 3,742<br>(51.44%) | 6,419<br>(46.63%) | 15,527<br>(45.40%) | 16,724<br>(44.84%) | 20,352<br>(45.04%) | 9,663<br>(43.58%) | 72,427<br>(45.29%) |
| Other | 0 | 0 | 2 (0.01%) | 0 | 0 | 0 | 2 (0.00%) |
| Unknown | 73 (1.00%) | 89 (0.65%) | 210 (0.61%) | 230 (0.62%) | 202 (0.45%) | 145 (0.65%) | 949<br>(0.59%) |
| Race |  |  |  |  |  |  |  |
| White | 3,382<br>(46.49%) | 7,278<br>(52.87%) | 19,256<br>(56.30%) | 21,559<br>(57.80%) | 30,094<br>(66.60%) | 14,472<br>(65.27%) | 96,041<br>(60.06%) |
| Black or African American | 1,406<br>(19.33%) | 2,308<br>(16.77%) | 6,148<br>(17.98%) | 7,553<br>(20.25%) | 6,839<br>(15.14%) | 3,764<br>(16.97%) | 28,018<br>(17.52%) |
| Asian | 411 (5.65%) | 761 (5.53%) | 1,542 (4.51%) | 1,156 (3.10%) | 1,134 (2.51%) | 652 (2.94%) | 5,656<br>(3.54%) |
| Native Hawaiian or Other Pacific Islander | 6 (0.08%) | 55 (0.40%) | 133 (0.39%) | 179 (0.48%) | 205 (0.45%) | 128 (0.58%) | 706<br>(0.44%) |
| American Indian or Alaska Native | 46 (0.63%) | 84 (0.61%) | 221 (0.65%) | 186 (0.50%) | 253 (0.56%) | 98 (0.44%) | 888<br>(0.56%) |
| Other Race | 1,300<br>(17.87%) | 1,901<br>(13.81%) | 3,380 (9.88%) | 2,928 (7.85%) | 2,881 (6.38%) | 1,250 (5.64%) | 13,640<br>(8.53%) |
| Unknown | 724 (9.95%) | 1,378<br>(10.01%) | 3,523<br>(10.30%) | 3,740<br>(10.03%) | 3,777 (8.36%) | 1,810 (8.16%) | 14,952<br>(9.35%) |
| Ethnicity |  |  |  |  |  |  |  |
| Hispanic or Latino | 875 (12.03%) | 1,664 (12.09%) | 3,765 (11.01%) | 3,914 (10.49%) | 5,006 (11.08%) | 2,270 (10.24%) | 17,494 (10.94%) |
| Not Hispanic or Latino | 5,259 (72.29%) | 10,558 (76.70%) | 26,950 (78.79%) | 29,843 (80.01%) | 36,910 (81.69%) | 18,431 (83.12%) | 127,951 (80.02%) |
| Unknown | 1,141 (15.68%) | 1,543 (11.21%) | 3,488 (10.20%) | 3,544 (9.50%) | 3,267 (7.23%) | 1,473 (6.64%) | 14,456 (9.04%) |
| Comorbidity |  |  |  |  |  |  |  |
| Asthma | 710 (9.76%) | 1,850 (13.44%) | 4,921 (14.39%) | 5,599 (15.01%) | 7,087 (15.69%) | 3,564 (16.07%) | 23,731 (14.84%) |
| Chronic Lung Disease | 225 (3.09%) | 779 (5.66%) | 2,358 (6.89%) | 2,855 (7.65%) | 4,472 (9.90%) | 2,178 (9.82%) | 12,867 (8.05%) |
| Chronic Obstructive Pulmonary Disease | 696 (9.57%) | 2,060 (14.97%) | 5,978 (17.48%) | 7,148 (19.16%) | 10,297 (22.79%) | 4,829 (21.78%) | 31,008 (19.39%) |

#### Hospitalization rate over time

The rate of influenza-associated hospitalization is shown in Figure 8. Figure 9 shows seasonal trends.

**Figure 8:**
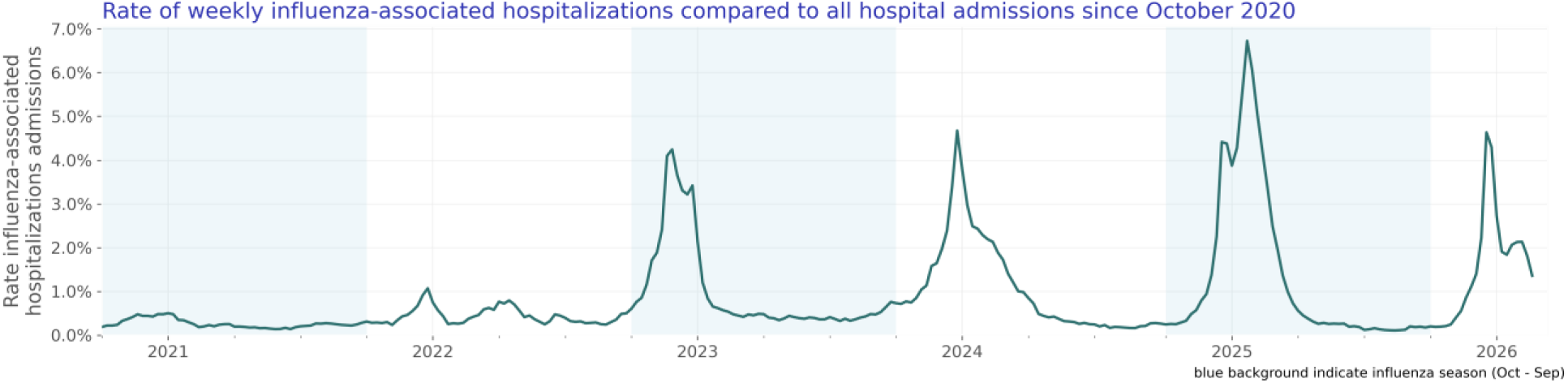
Rate of weekly influenza-associated hospitalizations compared to all hospital admissions since October 2020.

**Figure 9:**
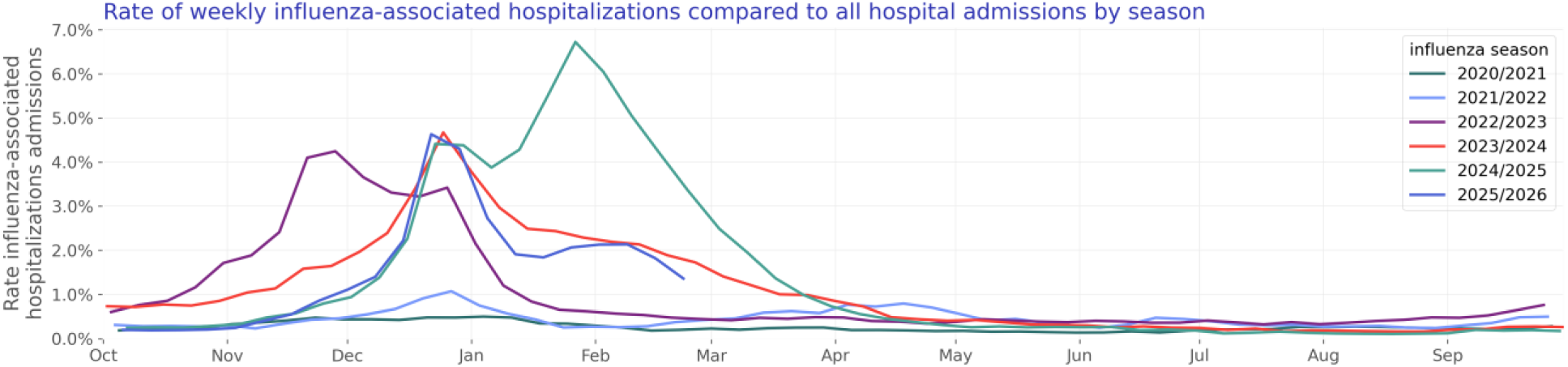
Rate of weekly influenza-associated hospitalizations compared to all hospital admissions by season.

#### Test positivity rate over time

We included 23,879,467 influenza lab results with known results of which 2,336,504 were positive. The influenza test positivity rate is shown in Figure 10. Figure 11 shows yearly trends.

**Figure 10:**
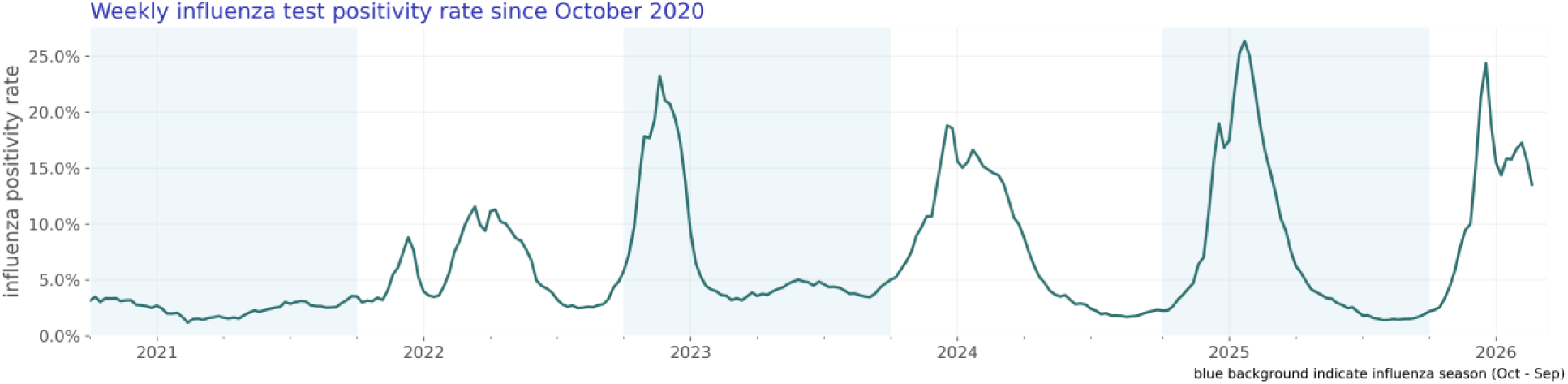
Weekly influenza test positivity rate since October 2020.

**Figure 11:**
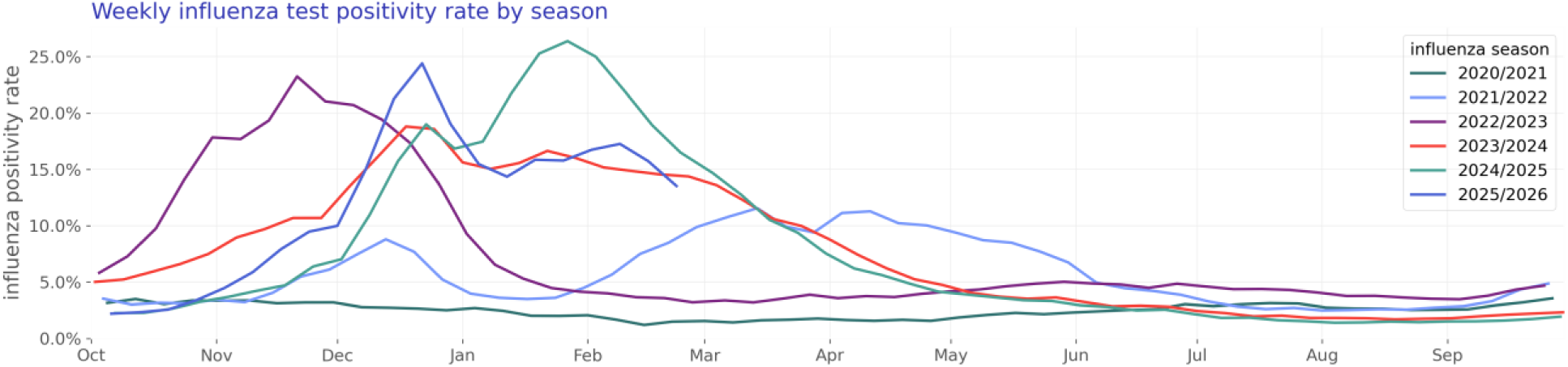
Weekly influenza test positivity rate by season.

### Human metapneumovirus (HMPV)

Our HMPV study population consists of 26,848 hospitalizations of 20,311 unique patients (Table 4).

**Table 4:** HMPV patient characteristics by season.

|  | 2020/2021 | 2021/2022 | 2022/2023 | 2023/2024 | 2024/2025 | 2025/2026 | Overall |
| --- | --- | --- | --- | --- | --- | --- | --- |
| N | 100 (100.00%) | 2,986 (100.00%) | 4,991 (100.00%) | 4,282 (100.00%) | 5,591 (100.00%) | 2,361 (100.00%) | 20,311 (100.00%) |
| Age Group |  |  |  |  |  |  |  |
| < 6 months | 1 (1.00%) | 81 (2.71%) | 180 (3.61%) | 95 (2.22%) | 115 (2.06%) | 41 (1.74%) | 513 (2.53%) |
| 6 - <12 months | 5 (5.00%) | 120 (4.02%) | 159 (3.19%) | 126 (2.94%) | 136 (2.43%) | 58 (2.46%) | 604 (2.97%) |
| 1 - <2 years | 10 (10.00%) | 170 (5.69%) | 203 (4.07%) | 114 (2.66%) | 158 (2.83%) | 70 (2.96%) | 725 (3.57%) |
| 2 - 4 years | 11 (11.00%) | 252 (8.44%) | 336 (6.73%) | 176 (4.11%) | 194 (3.47%) | 75 (3.18%) | 1,044 (5.14%) |
| 5 - 17 years | 1 (1.00%) | 131 (4.39%) | 189 (3.79%) | 138 (3.22%) | 147 (2.63%) | 60 (2.54%) | 666<br>(3.28%) |
| 18 - 49 years | 17 (17.00%) | 371 (12.42%) | 550 (11.02%) | 535 (12.49%) | 686 (12.27%) | 276 (11.69%) | 2,435<br>(11.99%) |
| 50 - 64 years | 24 (24.00%) | 551 (18.45%) | 869 (17.41%) | 775 (18.10%) | 948 (16.96%) | 388 (16.43%) | 3,555<br>(17.50%) |
| 65 - 74 years | 8 (8.00%) | 532 (17.82%) | 935 (18.73%) | 839 (19.59%) | 1,115<br>(19.94%) | 478 (20.25%) | 3,907<br>(19.24%) |
| 75 - 84 years | 15 (15.00%) | 501 (16.78%) | 964 (19.31%) | 933 (21.79%) | 1,271<br>(22.73%) | 518 (21.94%) | 4,202<br>(20.69%) |
| 85+ years | 8 (8.00%) | 277 (9.28%) | 606 (12.14%) | 551 (12.87%) | 821 (14.68%) | 397 (16.81%) | 2,660<br>(13.10%) |
| Sex |  |  |  |  |  |  |  |
| Female | 49 (49.00%) | 1,683<br>(56.36%) | 2,911<br>(58.32%) | 2,605<br>(60.84%) | 3,260<br>(58.31%) | 1,370<br>(58.03%) | 11,878<br>(58.48%) |
| Male | 49 (49.00%) | 1,282<br>(42.93%) | 2,064<br>(41.35%) | 1,658<br>(38.72%) | 2,309<br>(41.30%) | 967 (40.96%) | 8,329<br>(41.01%) |
| Unknown | 2 (2.00%) | 21 (0.70%) | 16 (0.32%) | 19 (0.44%) | 22 (0.39%) | 24 (1.02%) | 104<br>(0.51%) |
| Race |  |  |  |  |  |  |  |
| White | 71 (71.00%) | 2,192<br>(73.41%) | 3,566<br>(71.45%) | 2,900<br>(67.73%) | 3,818<br>(68.29%) | 1,672<br>(70.82%) | 14,219<br>(70.01%) |
| Black or African American | 7 (7.00%) | 329 (11.02%) | 519 (10.40%) | 511 (11.93%) | 712 (12.73%) | 242 (10.25%) | 2,320<br>(11.42%) |
| Asian | 0 | 56 (1.88%) | 134 (2.68%) | 99 (2.31%) | 147 (2.63%) | 70 (2.96%) | 506<br>(2.49%) |
| Native Hawaiian or Other Pacific Islander | 0 | 17 (0.57%) | 15 (0.30%) | 12 (0.28%) | 19 (0.34%) | 5 (0.21%) | 68<br>(0.33%) |
| American Indian or Alaska Native | 2 (2.00%) | 20 (0.67%) | 40 (0.80%) | 23 (0.54%) | 30 (0.54%) | 20 (0.85%) | 135<br>(0.66%) |
| Other Race | 10 (10.00%) | 162 (5.43%) | 333 (6.67%) | 316 (7.38%) | 377 (6.74%) | 142 (6.01%) | 1,340<br>(6.60%) |
| Unknown | 10 (10.00%) | 210 (7.03%) | 384 (7.69%) | 421 (9.83%) | 488 (8.73%) | 210 (8.89%) | 1,723<br>(8.48%) |
| Ethnicity |  |  |  |  |  |  |  |
| Hispanic or Latino | 22 (22.00%) | 347 (11.62%) | 674 (13.50%) | 636 (14.85%) | 759 (13.58%) | 305 (12.92%) | 2,743<br>(13.50%) |
| Not Hispanic or Latino | 70 (70.00%) | 2,410<br>(80.71%) | 3,979<br>(79.72%) | 3,317<br>(77.46%) | 4,428<br>(79.20%) | 1,902<br>(80.56%) | 16,106<br>(79.30%) |
| Unknown | 8 (8.00%) | 229 (7.67%) | 338 (6.77%) | 329 (7.68%) | 404 (7.23%) | 154 (6.52%) | 1,462<br>(7.20%) |
| Comorbidity |  |  |  |  |  |  |  |
| Asthma | 13 (13.00%) | 442 (14.80%) | 811 (16.25%) | 760 (17.75%) | 971 (17.37%) | 427 (18.09%) | 3,424<br>(16.86%) |
| Chronic Lung Disease | 12 (12.00%) | 334 (11.19%) | 505 (10.12%) | 433 (10.11%) | 638 (11.41%) | 256 (10.84%) | 2,178<br>(10.72%) |
| Chronic Obstructive Pulmonary Disease | 17 (17.00%) | 606 (20.29%) | 1,033<br>(20.70%) | 874 (20.41%) | 1,240<br>(22.18%) | 490 (20.75%) | 4,260<br>(20.97%) |

#### Hospitalization rate over time

The rate of HMPV-associated hospitalization is shown in Figure 12. Figure 13 shows seasonal trends.

**Figure 12:**
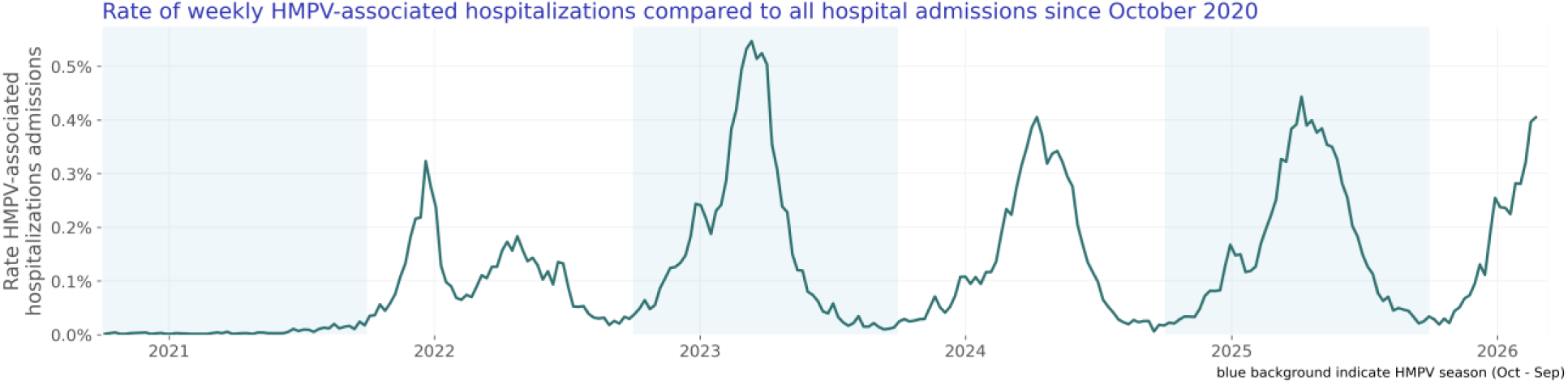
Rate of weekly HMPV-associated hospitalizations compared to all hospital admissions since October 2020.

**Figure 13:**
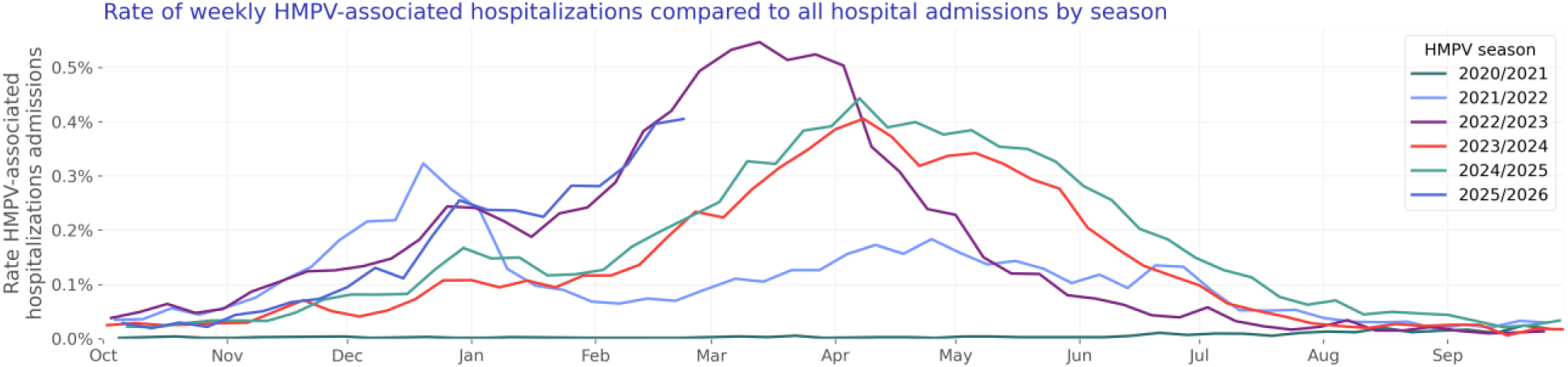
Rate of weekly HMPV-associated hospitalizations compared to all hospital admissions by season.

#### Test positivity rate over time

We included 2,742,681 HMPV lab results with known results of which 70,775 were positive. The HMPV test positivity rate is shown in Figure 14. Figure 15 shows yearly trends.

**Figure 14:**
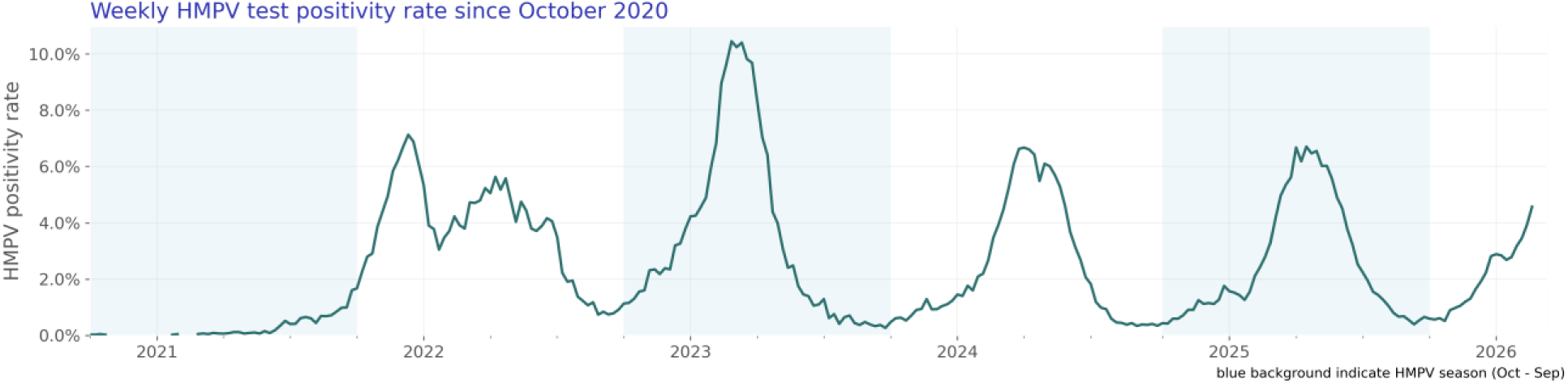
Weekly HMPV test positivity rate since October 2020.

**Figure 15:**
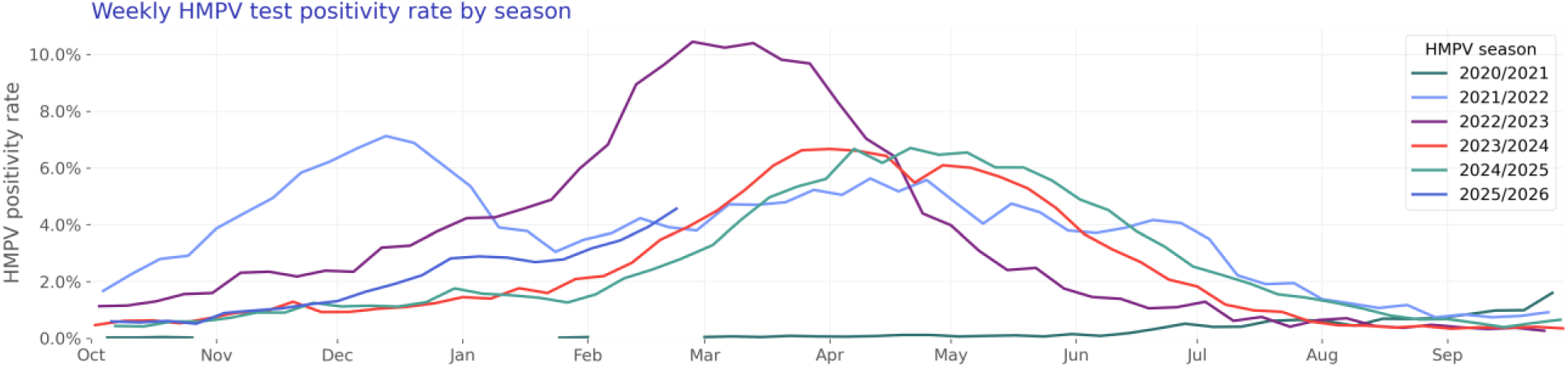
Weekly HMPV test positivity rate by season.

### Parainfluenza virus

Our parainfluenza study population consists of 28,613 hospitalizations of 21,108 unique patients (Table 5).

**Table 5:**
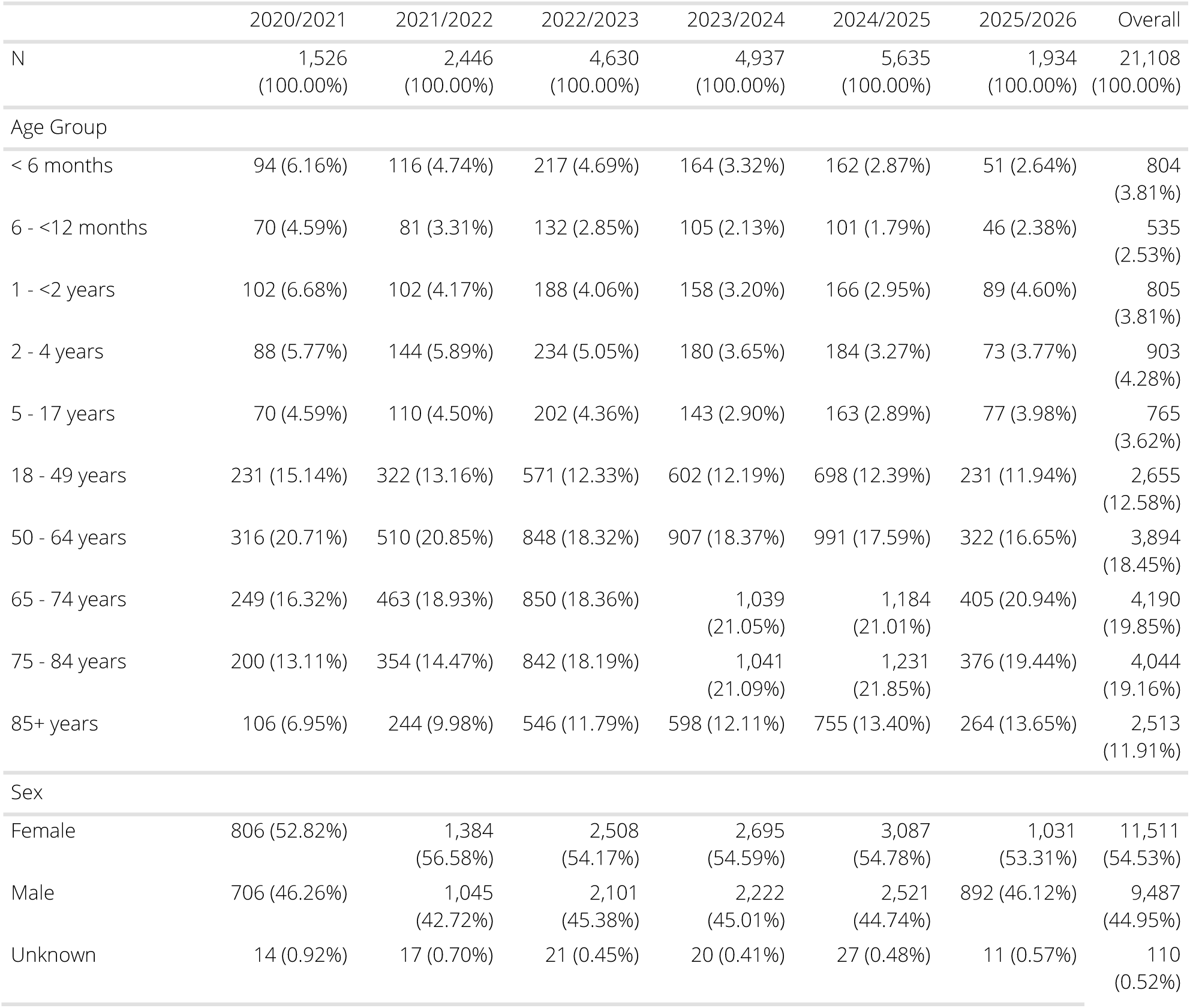
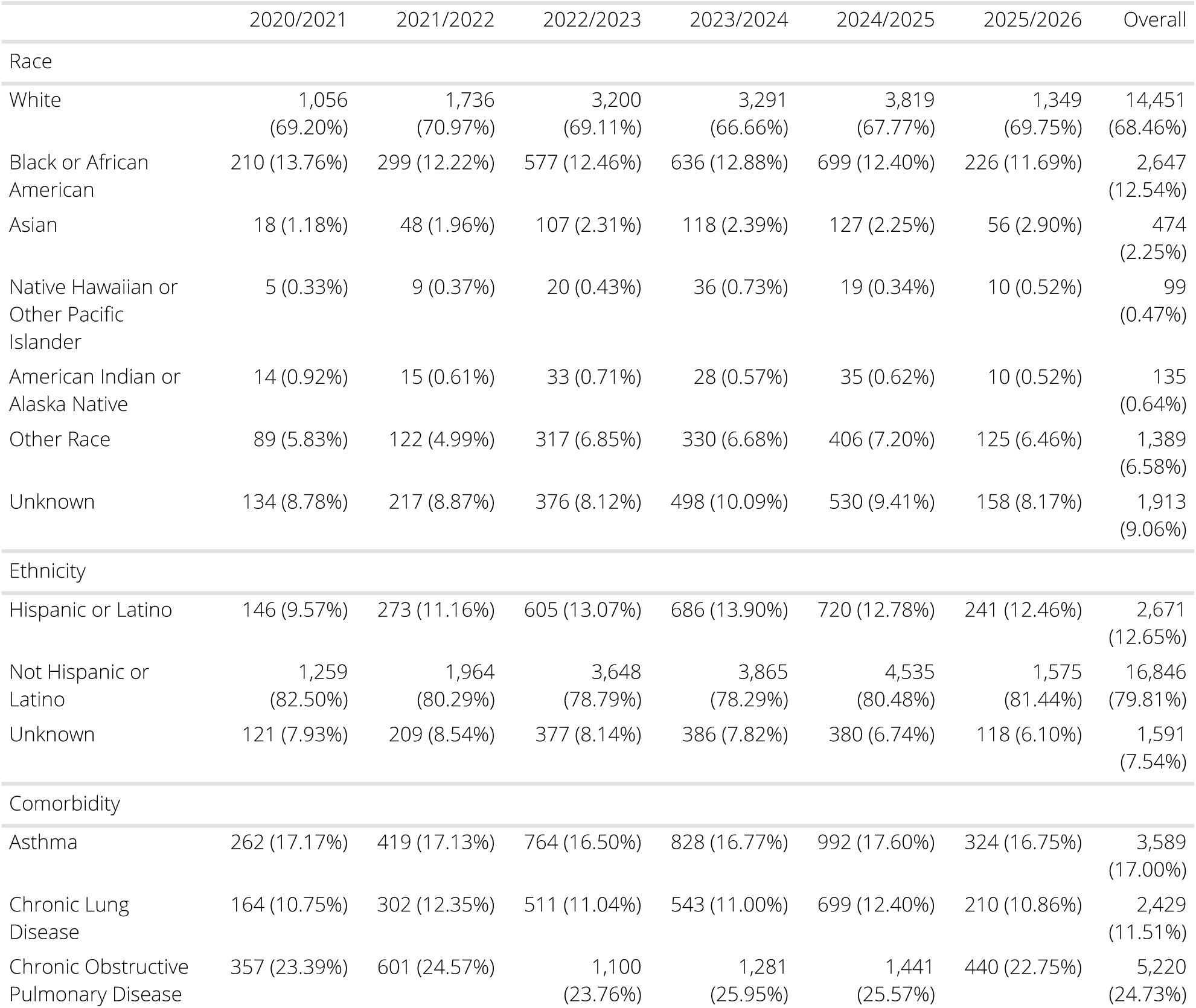
Parainfluenza patient characteristics by season.

#### Hospitalization rate over time

The rate of parainfluenza-associated hospitalization is shown in Figure 16. Figure 17 shows seasonal trends.

**Figure 16:**
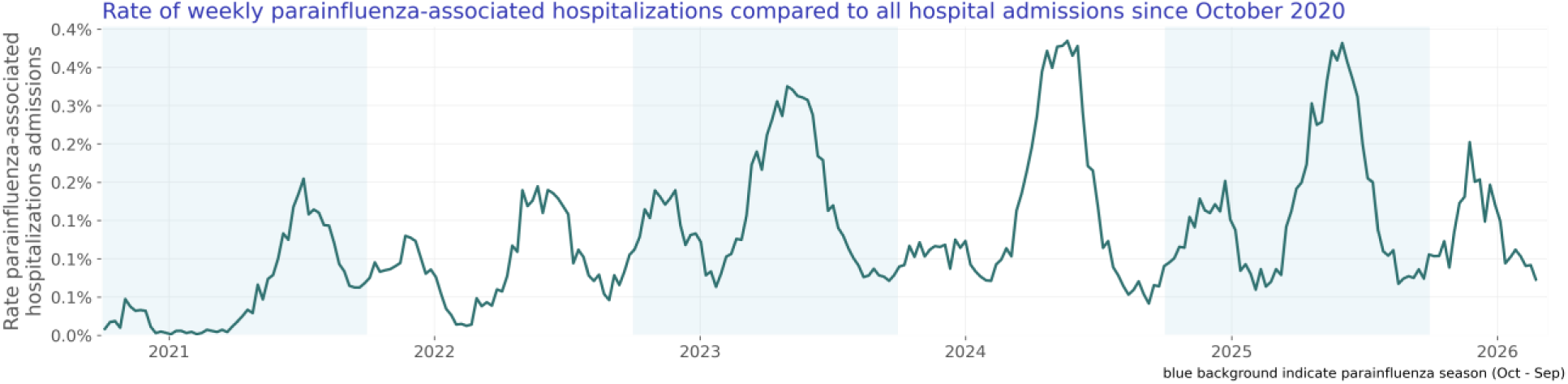
Rate of weekly parainfluenza-associated hospitalizations compared to all hospital admissions since October 2020.

**Figure 17:**
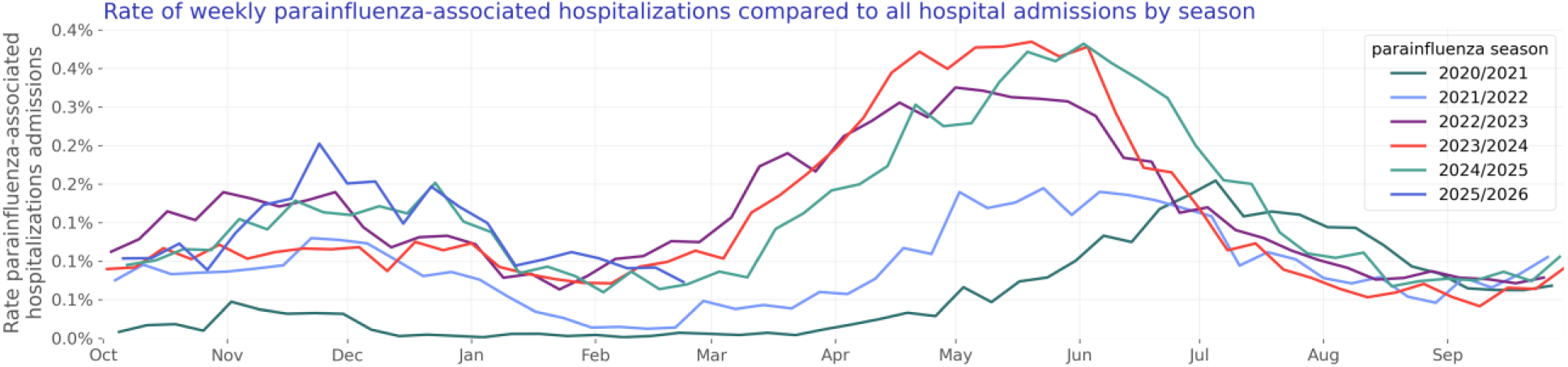
Rate of weekly parainfluenza-associated hospitalizations compared to all hospital admissions by season.

#### Test positivity rate over time

We included 2,870,366 parainfluenza lab results with known results of which 107,410 were positive. The parainfluenza test positivity rate is shown in Figure 18. Figure 19 shows yearly trends.

**Figure 18:**
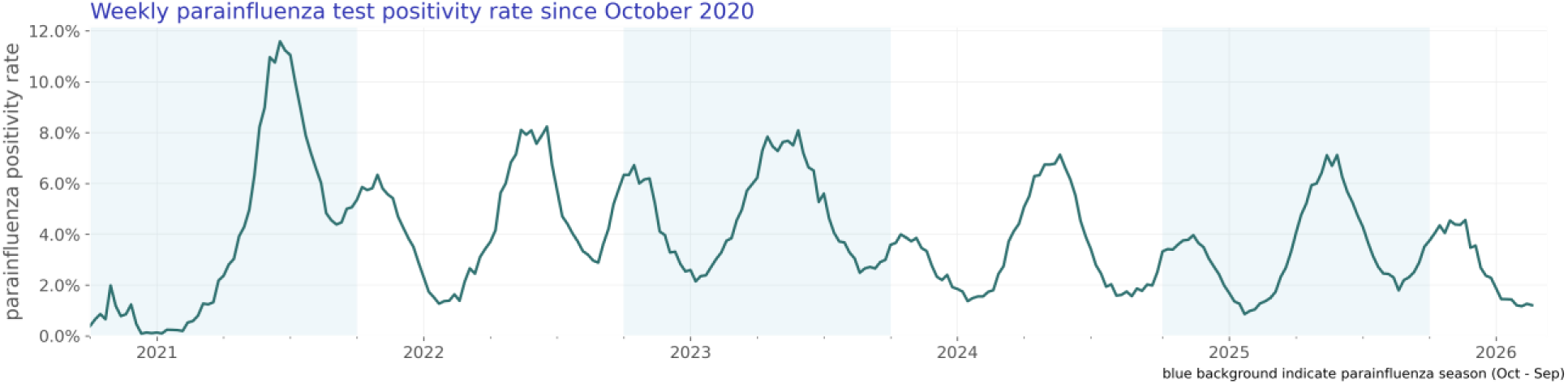
Weekly parainfluenza test positivity rate since October 2020.

**Figure 19:**
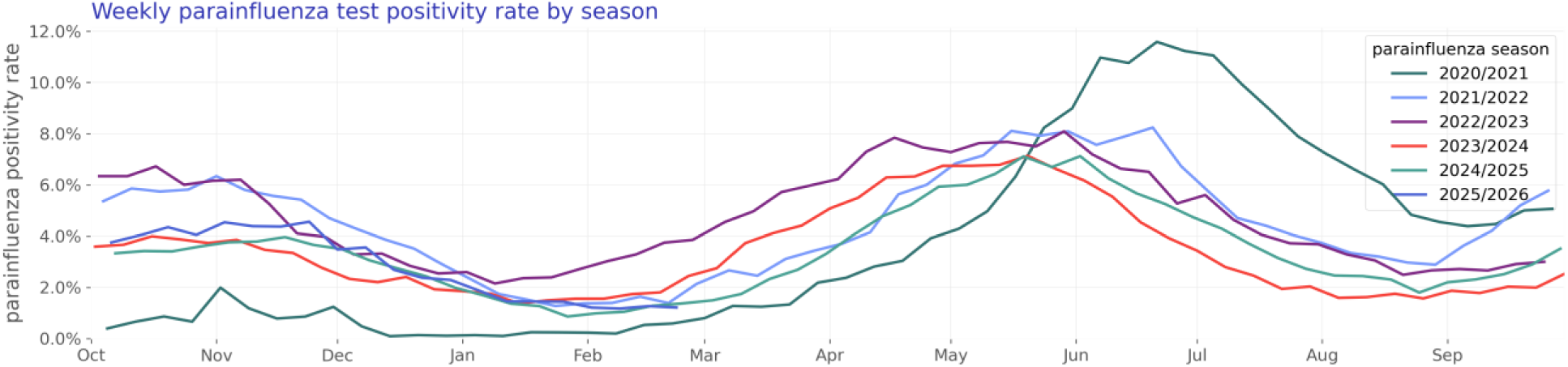
Weekly parainfluenza test positivity rate by season.

### Respiratory syncytial virus (RSV)

Our RSV study population consists of 65,247 hospitalizations of 47,925 unique patients (Table 6).

**Table 6:** RSV patient characteristics by season.

|  | 2020/2021 | 2021/2022 | 2022/2023 | 2023/2024 | 2024/2025 | 2025/2026 | Overall |
| --- | --- | --- | --- | --- | --- | --- | --- |
| N | 2,339<br>(100.00%) | 4,955<br>(100.00%) | 9,513<br>(100.00%) | 11,910<br>(100.00%) | 12,774<br>(100.00%) | 6,434<br>(100.00%) | 47,925<br>(100.00%) |
| Age Group |  |  |  |  |  |  |  |
| < 6 months | 600 (25.65%) | 900 (18.16%) | 1,689<br>(17.75%) | 1,292<br>(10.85%) | 966 (7.56%) | 448 (6.96%) | 5,895<br>(12.30%) |
| 6 - <12 months | 202 (8.64%) | 375 (7.57%) | 671 (7.05%) | 607 (5.10%) | 593 (4.64%) | 277 (4.31%) | 2,725<br>(5.69%) |
| 1 - <2 years | 232 (9.92%) | 415 (8.38%) | 741 (7.79%) | 635 (5.33%) | 683 (5.35%) | 452 (7.03%) | 3,158<br>(6.59%) |
| 2 - 4 years | 190 (8.12%) | 420 (8.48%) | 826 (8.68%) | 527 (4.42%) | 705 (5.52%) | 380 (5.91%) | 3,048<br>(6.36%) |
| 5 - 17 years | 45 (1.92%) | 130 (2.62%) | 290 (3.05%) | 257 (2.16%) | 299 (2.34%) | 153 (2.38%) | 1,174<br>(2.45%) |
| 18 - 49 years | 232 (9.92%) | 440 (8.88%) | 798 (8.39%) | 1,069 (8.98%) | 1,269 (9.93%) | 676 (10.51%) | 4,484<br>(9.36%) |
| 50 - 64 years | 280 (11.97%) | 608 (12.27%) | 1,087<br>(11.43%) | 1,760<br>(14.78%) | 1,933<br>(15.13%) | 900 (13.99%) | 6,568<br>(13.70%) |
| 65 - 74 years | 255 (10.90%) | 619 (12.49%) | 1,254<br>(13.18%) | 2,007<br>(16.85%) | 2,192<br>(17.16%) | 1,083<br>(16.83%) | 7,410<br>(15.46%) |
| 75 - 84 years | 193 (8.25%) | 657 (13.26%) | 1,282<br>(13.48%) | 2,196<br>(18.44%) | 2,460<br>(19.26%) | 1,167<br>(18.14%) | 7,955<br>(16.60%) |
| 85+ years | 110 (4.70%) | 391 (7.89%) | 875 (9.20%) | 1,560<br>(13.10%) | 1,674<br>(13.10%) | 898 (13.96%) | 5,508<br>(11.49%) |
| Sex |  |  |  |  |  |  |  |
| Female | 1,282<br>(54.81%) | 2,546<br>(51.38%) | 5,063<br>(53.22%) | 6,538<br>(54.90%) | 6,967<br>(54.54%) | 3,524<br>(54.77%) | 25,920<br>(54.08%) |
| Male | 1,049<br>(44.85%) | 2,387<br>(48.17%) | 4,405<br>(46.31%) | 5,316<br>(44.63%) | 5,750<br>(45.01%) | 2,864<br>(44.51%) | 21,771<br>(45.43%) |
| Unknown | 8 (0.34%) | 22 (0.44%) | 45 (0.47%) | 56 (0.47%) | 57 (0.45%) | 46 (0.71%) | 234<br>(0.49%) |
| Race |  |  |  |  |  |  |  |
| White | 1,517<br>(64.86%) | 3,413<br>(68.88%) | 6,322<br>(66.46%) | 8,167<br>(68.57%) | 8,256<br>(64.63%) | 4,107<br>(63.83%) | 31,782<br>(66.32%) |
| Black or African American | 420 (17.96%) | 609 (12.29%) | 1,165<br>(12.25%) | 1,490<br>(12.51%) | 1,760<br>(13.78%) | 998 (15.51%) | 6,442<br>(13.44%) |
| Asian | 37 (1.58%) | 141 (2.85%) | 314 (3.30%) | 305 (2.56%) | 358 (2.80%) | 215 (3.34%) | 1,370<br>(2.86%) |
| Native Hawaiian or Other Pacific Islander | 16 (0.68%) | 70 (1.41%) | 154 (1.62%) | 108 (0.91%) | 171 (1.34%) | 90 (1.40%) | 609<br>(1.27%) |
| American Indian or Alaska Native | 16 (0.68%) | 34 (0.69%) | 64 (0.67%) | 60 (0.50%) | 69 (0.54%) | 28 (0.44%) | 271<br>(0.57%) |
| Other Race | 160 (6.84%) | 265 (5.35%) | 592 (6.22%) | 603 (5.06%) | 956 (7.48%) | 421 (6.54%) | 2,997<br>(6.25%) |
| Unknown | 173 (7.40%) | 423 (8.54%) | 902 (9.48%) | 1,177 (9.88%) | 1,204 (9.43%) | 575 (8.94%) | 4,454<br>(9.29%) |
| Ethnicity |  |  |  |  |  |  |  |
| Hispanic or Latino | 336 (14.37%) | 669 (13.50%) | 1,391<br>(14.62%) | 1,453<br>(12.20%) | 1,861<br>(14.57%) | 883 (13.72%) | 6,593<br>(13.76%) |
| Not Hispanic or Latino | 1,822<br>(77.90%) | 3,890<br>(78.51%) | 7,363<br>(77.40%) | 9,519<br>(79.92%) | 9,967<br>(78.03%) | 5,132<br>(79.76%) | 37,693<br>(78.65%) |
| Unknown | 181 (7.74%) | 396 (7.99%) | 759 (7.98%) | 938 (7.88%) | 946 (7.41%) | 419 (6.51%) | 3,639<br>(7.59%) |
| Comorbidity |  |  |  |  |  |  |  |
| Asthma | 280 (11.97%) | 616 (12.43%) | 1,270<br>(13.35%) | 1,758<br>(14.76%) | 2,121<br>(16.60%) | 1,093<br>(16.99%) | 7,138<br>(14.89%) |
| Chronic Lung Disease | 150 (6.41%) | 346 (6.98%) | 667 (7.01%) | 985 (8.27%) | 1,223 (9.57%) | 578 (8.98%) | 3,949<br>(8.24%) |
| Chronic Obstructive Pulmonary Disease | 318 (13.60%) | 813 (16.41%) | 1,575<br>(16.56%) | 2,518<br>(21.14%) | 2,739<br>(21.44%) | 1,286<br>(19.99%) | 9,249<br>(19.30%) |

#### Hospitalization rate over time

The rate of RSV-associated hospitalization is shown in Figure 20. Figure 21 shows seasonal trends.

**Figure 20:**
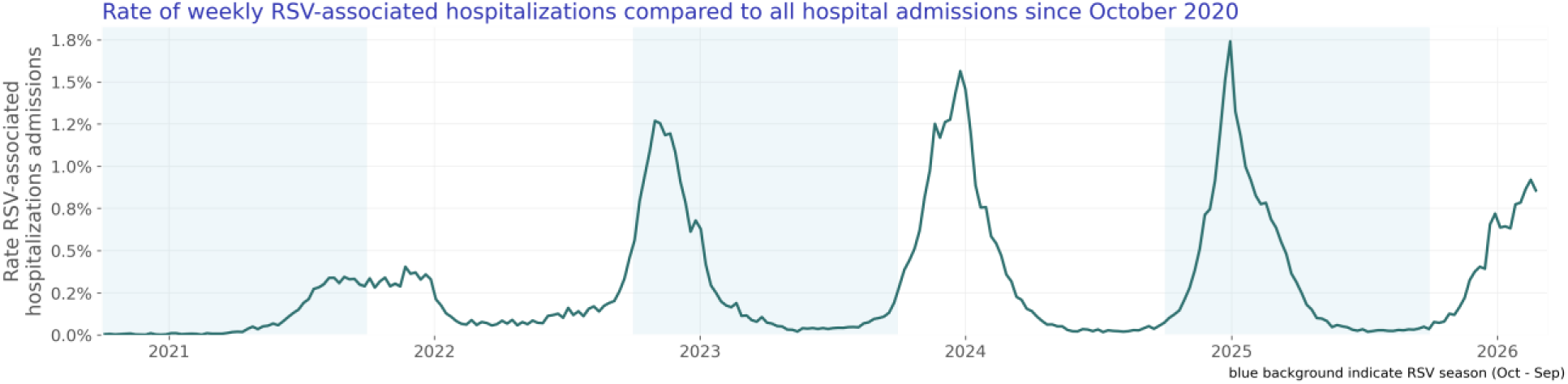
Rate of weekly RSV-associated hospitalizations compared to all hospital admissions since October 2020.

**Figure 21:**
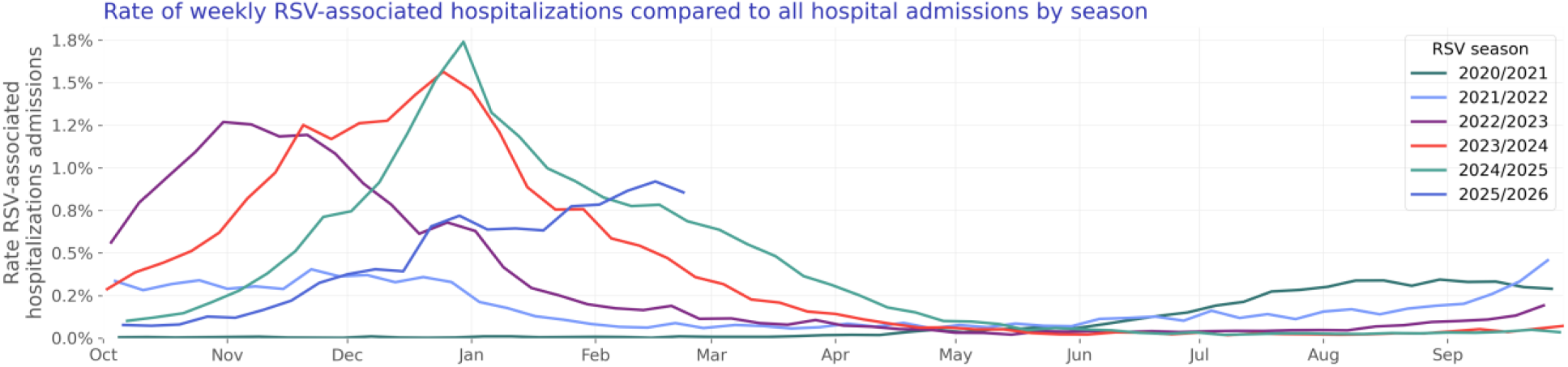
Rate of weekly RSV-associated hospitalizations compared to all hospital admissions by season.

#### Test positivity rate over time

We included 9,930,524 RSV lab results with known results of which 462,170 were positive. The RSV test positivity rate is shown in Figure 22. Figure 23 shows yearly trends.

**Figure 22:**
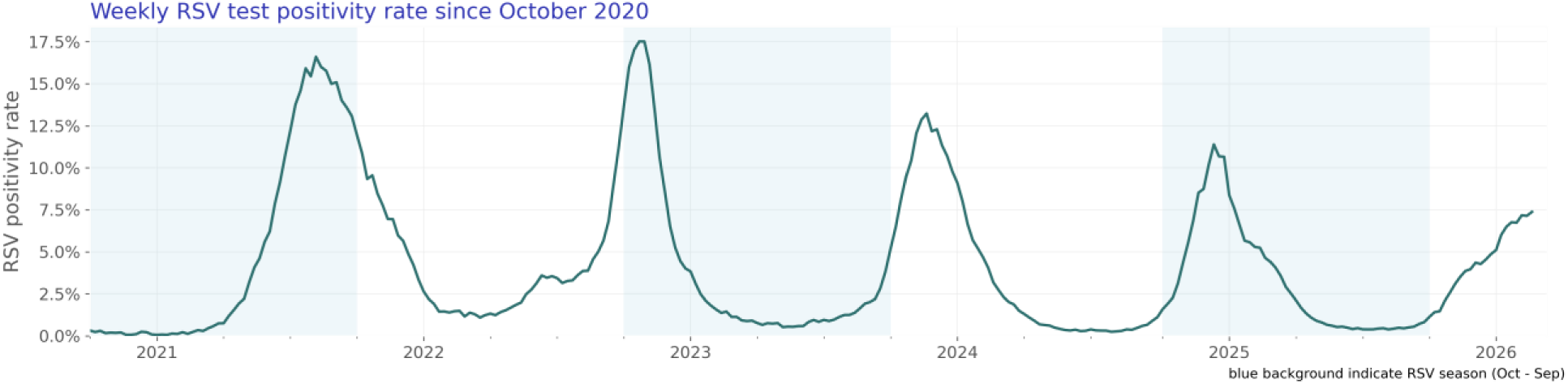
Weekly RSV test positivity rate since October 2020.

**Figure 23:**
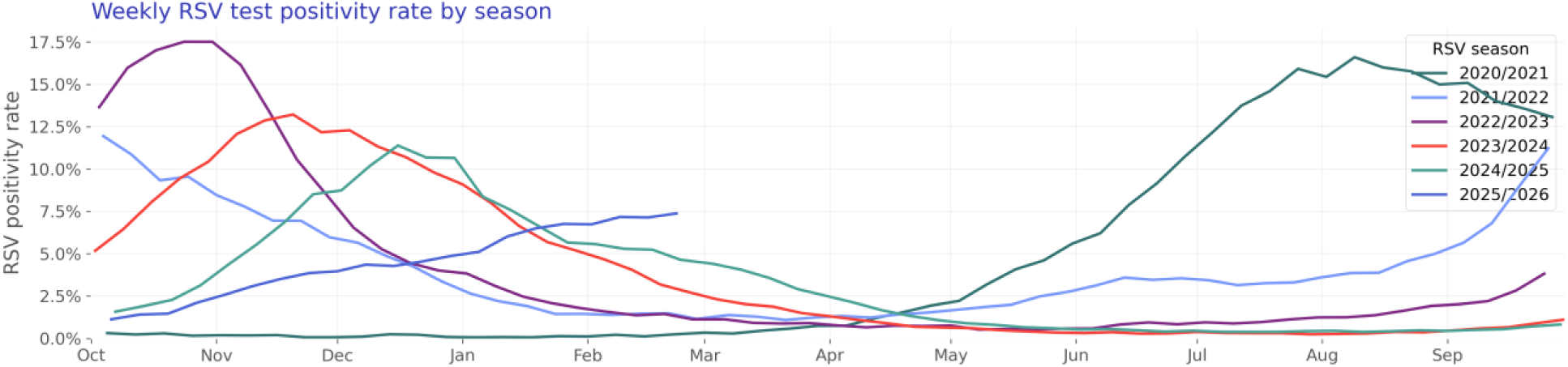
Weekly RSV test positivity rate by season.

### Rhinovirus

Our rhinovirus study population consists of 101,002 hospitalizations of 68,573 unique patients (Table 7).

**Table 7:**
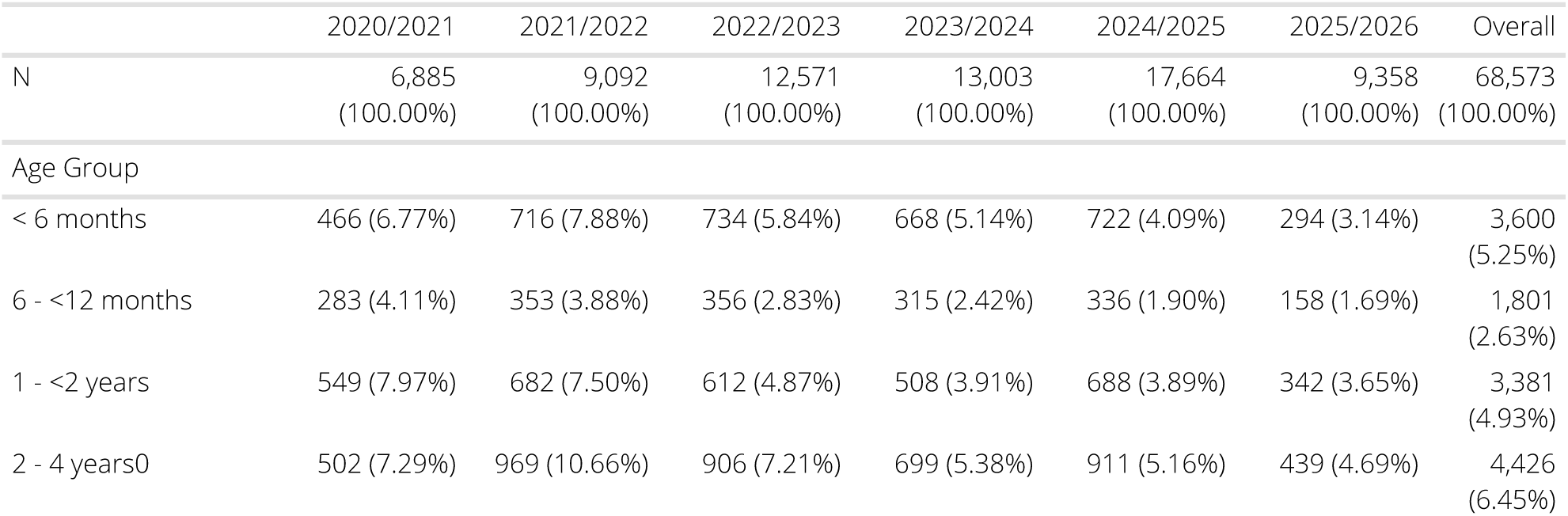
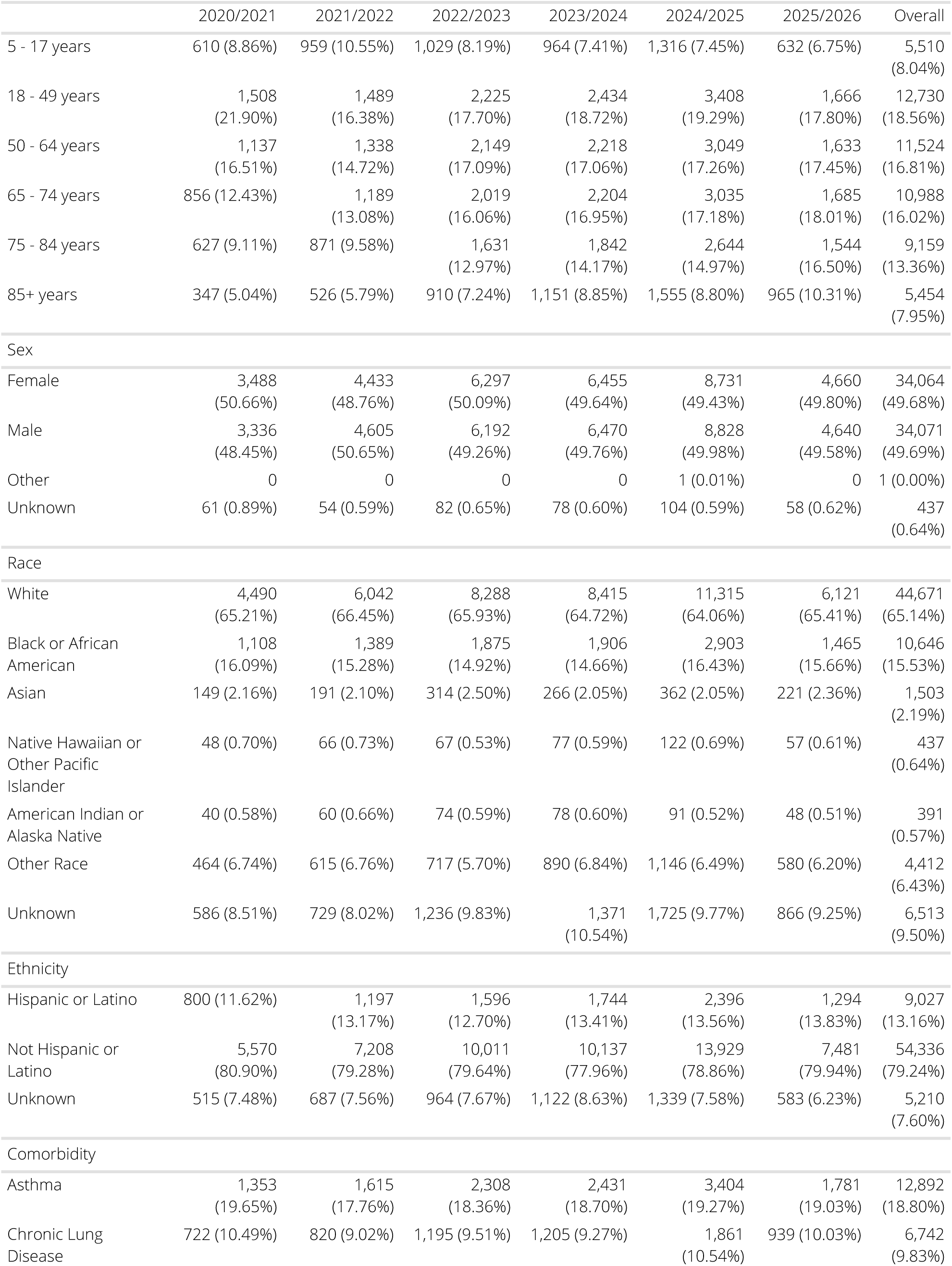
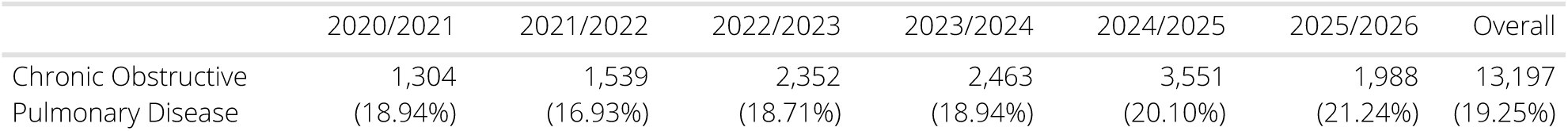
Rhinovirus patient characteristics by season.

#### Hospitalization rate over time

The rate of rhinovirus-associated hospitalization is shown in Figure 24. Figure 25 shows seasonal trends.

**Figure 24:**
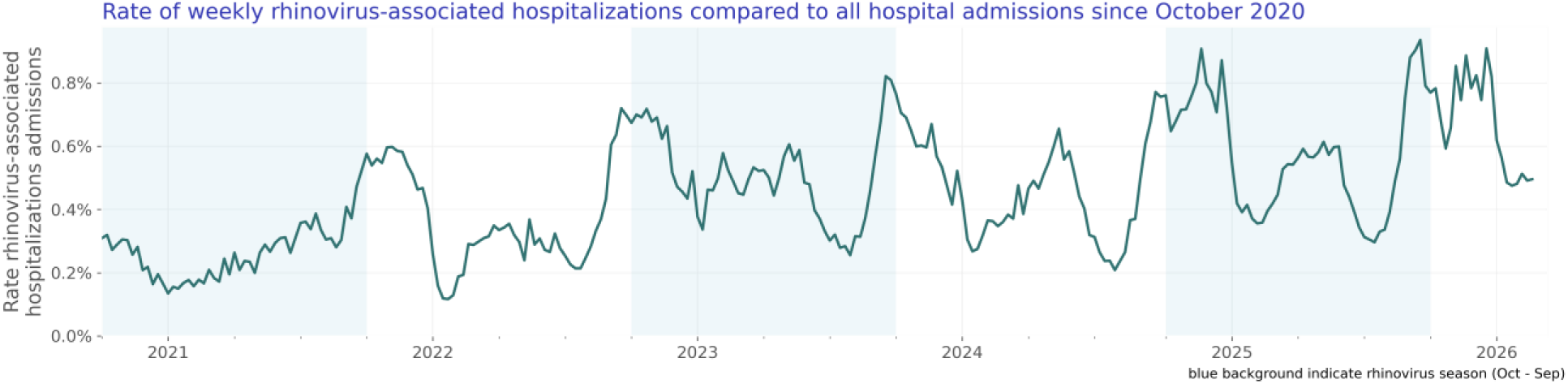
Rate of weekly rhinovirus-associated hospitalizations compared to all hospital admissions since October 2020.

**Figure 25:**
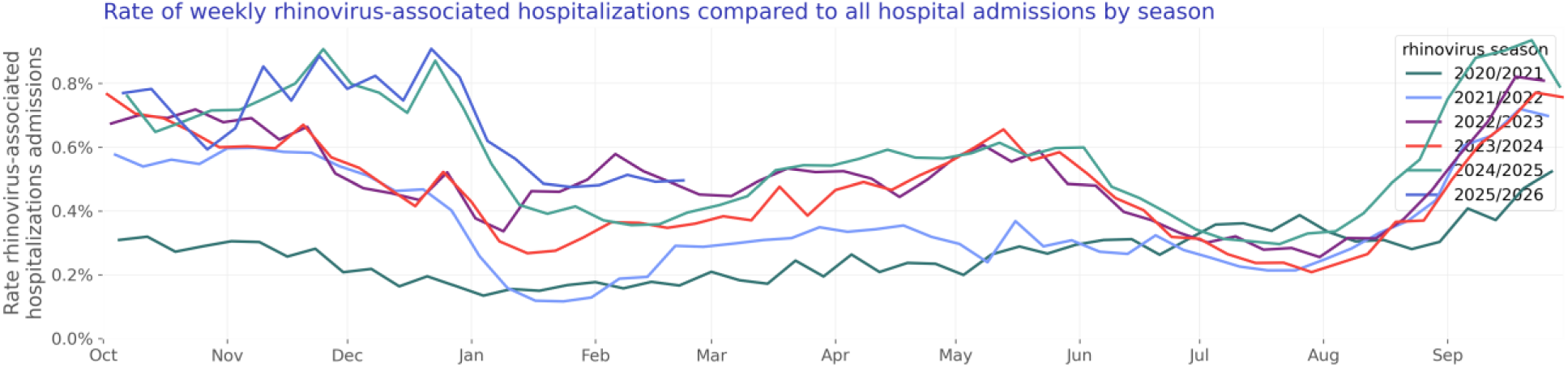
Rate of weekly rhinovirus-associated hospitalizations compared to all hospital admissions by season.

#### Test positivity rate over time

We included 2,300,813 rhinovirus lab results with known results of which 353,831 were positive. The rhinovirus test positivity rate is shown in Figure 26. Figure 27 shows yearly trends.

**Figure 26:**
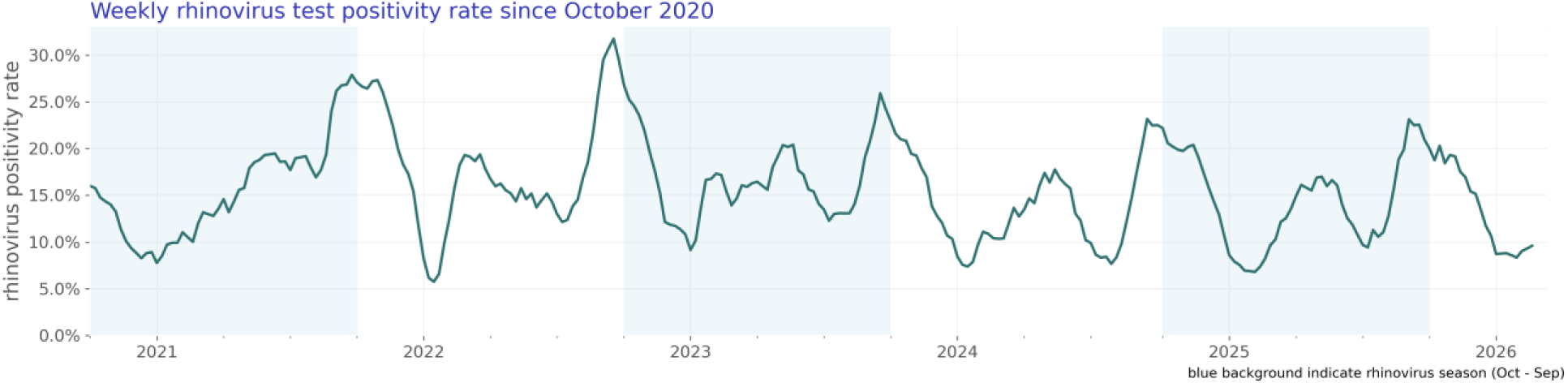
Weekly rhinovirus test positivity rate since October 2020.

**Figure 27:**
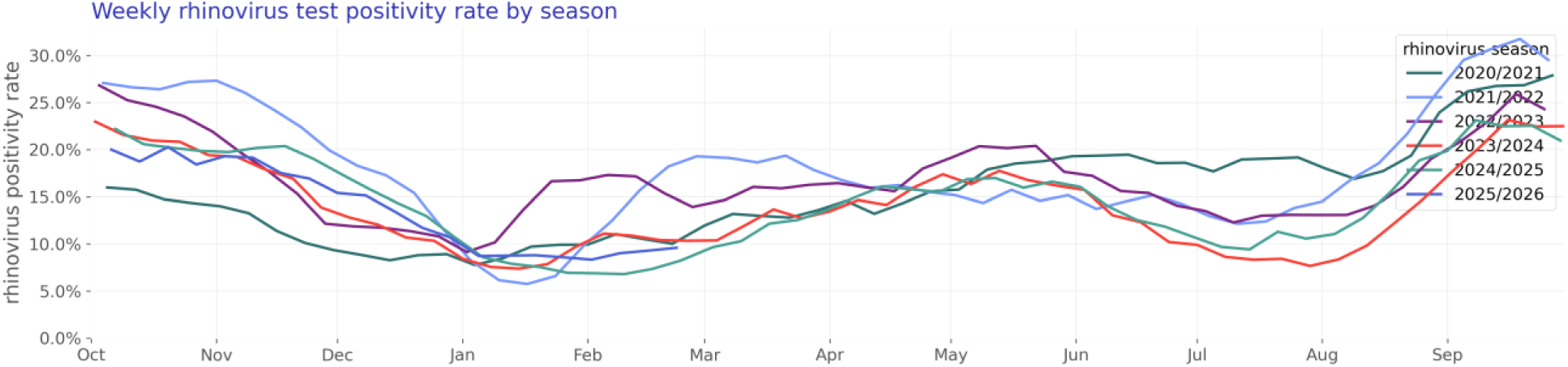
Weekly rhinovirus test positivity rate by season.

### Infants and children (age 0-4)

Our infants and children study population consists of 74,495 hospitalizations of 50,580 unique patients (Table 8).

**Table 8:** Infants and children patient characteristics by season.

|  | 2020/2021 | 2021/2022 | 2022/2023 | 2023/2024 | 2024/2025 | 2025/2026 | Overall |
| --- | --- | --- | --- | --- | --- | --- | --- |
| N | 5,282<br>(100.00%) | 9,891<br>(100.00%) | 12,319<br>(100.00%) | 9,566<br>(100.00%) | 9,216<br>(100.00%) | 4,306<br>(100.00%) | 50,580<br>(100.00%) |
| Age Group |  |  |  |  |  |  |  |
| < 6 months | 1,792<br>(33.93%) | 3,133<br>(31.68%) | 4,021<br>(32.64%) | 3,177<br>(33.21%) | 2,478<br>(26.89%) | 1,025<br>(23.80%) | 15,626<br>(30.89%) |
| 6 - <12 months | 785 (14.86%) | 1,506<br>(15.23%) | 1,963<br>(15.93%) | 1,693<br>(17.70%) | 1,517<br>(16.46%) | 680 (15.79%) | 8,144<br>(16.10%) |
| 1 - <2 years | 1,305<br>(24.71%) | 2,131<br>(21.54%) | 2,590<br>(21.02%) | 2,111<br>(22.07%) | 2,315<br>(25.12%) | 1,195<br>(27.75%) | 11,647<br>(23.03%) |
| 2 - 4 years | 1,400<br>(26.51%) | 3,121<br>(31.55%) | 3,745<br>(30.40%) | 2,585<br>(27.02%) | 2,906<br>(31.53%) | 1,406<br>(32.65%) | 15,163<br>(29.98%) |
| Sex |  |  |  |  |  |  |  |
| Female | 2,248<br>(42.56%) | 4,125<br>(41.70%) | 5,328<br>(43.25%) | 4,150<br>(43.38%) | 4,014<br>(43.55%) | 1,849<br>(42.94%) | 21,714<br>(42.93%) |
| Male | 3,034<br>(57.44%) | 5,764<br>(58.28%) | 6,990<br>(56.74%) | 5,414<br>(56.60%) | 5,200<br>(56.42%) | 2,457<br>(57.06%) | 28,859<br>(57.06%) |
| Unknown | 0 | 2 (0.02%) | 1 (0.01%) | 2 (0.02%) | 2 (0.02%) | 0 | 7 (0.01%) |
| Race |  |  |  |  |  |  |  |
| White | 2,740<br>(51.87%) | 5,179<br>(52.36%) | 6,102<br>(49.53%) | 4,743<br>(49.58%) | 4,551<br>(49.38%) | 2,263<br>(52.55%) | 25,578<br>(50.57%) |
| Black or African American | 972 (18.40%) | 1,600<br>(16.18%) | 1,919<br>(15.58%) | 1,483<br>(15.50%) | 1,455<br>(15.79%) | 715 (16.60%) | 8,144<br>(16.10%) |
| Asian | 225 (4.26%) | 563 (5.69%) | 875 (7.10%) | 542 (5.67%) | 453 (4.92%) | 218 (5.06%) | 2,876<br>(5.69%) |
| Native Hawaiian or Other Pacific Islander | 73 (1.38%) | 174 (1.76%) | 221 (1.79%) | 205 (2.14%) | 250 (2.71%) | 118 (2.74%) | 1,041<br>(2.06%) |
| American Indian or Alaska Native | 35 (0.66%) | 75 (0.76%) | 124 (1.01%) | 62 (0.65%) | 60 (0.65%) | 23 (0.53%) | 379<br>(0.75%) |
| Other Race | 905 (17.13%) | 1,491<br>(15.07%) | 1,878<br>(15.24%) | 1,435<br>(15.00%) | 1,501<br>(16.29%) | 563 (13.07%) | 7,773<br>(15.37%) |
| Unknown | 332 (6.29%) | 809 (8.18%) | 1,200 (9.74%) | 1,096<br>(11.46%) | 946 (10.26%) | 406 (9.43%) | 4,789<br>(9.47%) |
| Ethnicity |  |  |  |  |  |  |  |
| Hispanic or Latino | 1,119<br>(21.19%) | 2,121<br>(21.44%) | 2,998<br>(24.34%) | 2,426<br>(25.36%) | 2,516<br>(27.30%) | 1,109<br>(25.75%) | 12,289<br>(24.30%) |
| Not Hispanic or Latino | 3,793<br>(71.81%) | 7,042<br>(71.20%) | 8,432<br>(68.45%) | 6,512<br>(68.07%) | 6,092<br>(66.10%) | 2,915<br>(67.70%) | 34,786<br>(68.77%) |
| Unknown | 370 (7.00%) | 728 (7.36%) | 889 (7.22%) | 628 (6.56%) | 608 (6.60%) | 282 (6.55%) | 3,505<br>(6.93%) |
| Virus |  |  |  |  |  |  |  |
| COVID | 626 (11.85%) | 1,723<br>(17.42%) | 865 (7.02%) | 1,184<br>(12.38%) | 1,696<br>(18.40%) | 555 (12.89%) | 6,649<br>(13.15%) |
| HMPV | 27 (0.51%) | 623 (6.30%) | 878 (7.13%) | 511 (5.34%) | 603 (6.54%) | 244 (5.67%) | 2,886<br>(5.71%) |
| RSV | 1,224<br>(23.17%) | 2,110<br>(21.33%) | 3,927<br>(31.88%) | 3,061<br>(32.00%) | 2,947<br>(31.98%) | 1,557<br>(36.16%) | 14,826<br>(29.31%) |
| influenza | 1,251<br>(23.68%) | 2,272<br>(22.97%) | 3,270<br>(26.54%) | 2,013<br>(21.04%) | 700 (7.60%) | 458 (10.64%) | 9,964<br>(19.70%) |
| parainfluenza | 354 (6.70%) | 443 (4.48%) | 771 (6.26%) | 607 (6.35%) | 613 (6.65%) | 259 (6.01%) | 3,047<br>(6.02%) |
| rhinovirus | 1,800<br>(34.08%) | 2,720<br>(27.50%) | 2,608<br>(21.17%) | 2,190<br>(22.89%) | 2,657<br>(28.83%) | 1,233<br>(28.63%) | 13,208<br>(26.11%) |
| Comorbidity |  |  |  |  |  |  |  |
| Asthma | 394 (7.46%) | 626 (6.33%) | 821 (6.66%) | 759 (7.93%) | 900 (9.77%) | 458 (10.64%) | 3,958<br>(7.83%) |
| Chronic Lung Disease | 35 (0.66%) | 57 (0.58%) | 80 (0.65%) | 67 (0.70%) | 70 (0.76%) | 37 (0.86%) | 346<br>(0.68%) |
| Chronic Obstructive Pulmonary Disease | 8 (0.15%) | 17 (0.17%) | 17 (0.14%) | 21 (0.22%) | 12 (0.13%) | 15 (0.35%) | 90<br>(0.18%) |

The rate of respiratory virus-associated hospitalizations compared to all hospitalizations for infants and children under five is shown in Figure 28. Figure 29 shows the same data stacked to represent the combined impact of the viruses.

**Figure 28:**
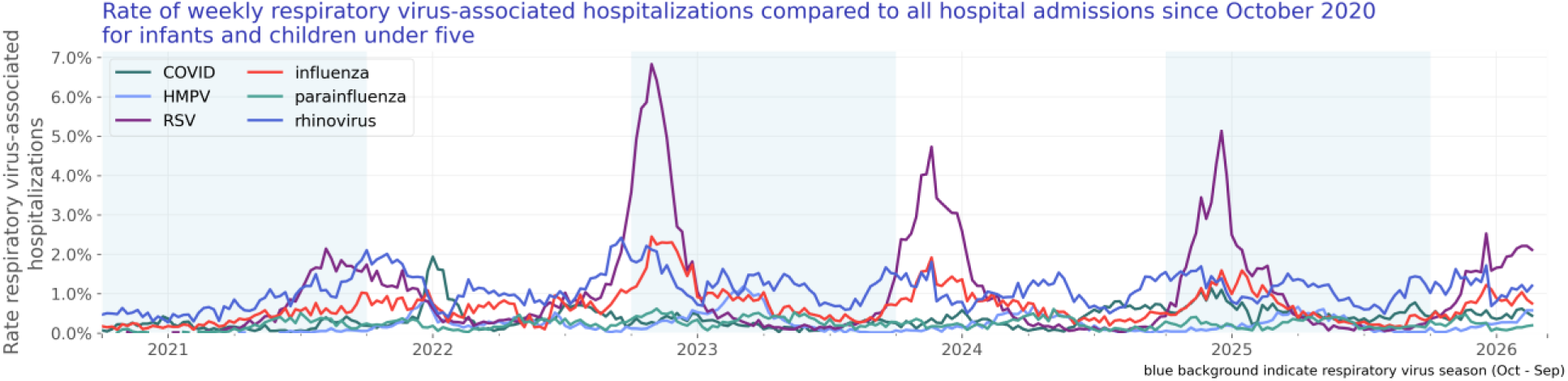
Rate of weekly respiratory virus-associated hospitalizations compared to all hospital admissions since October 2020 for infants and children under five.

**Figure 29:**
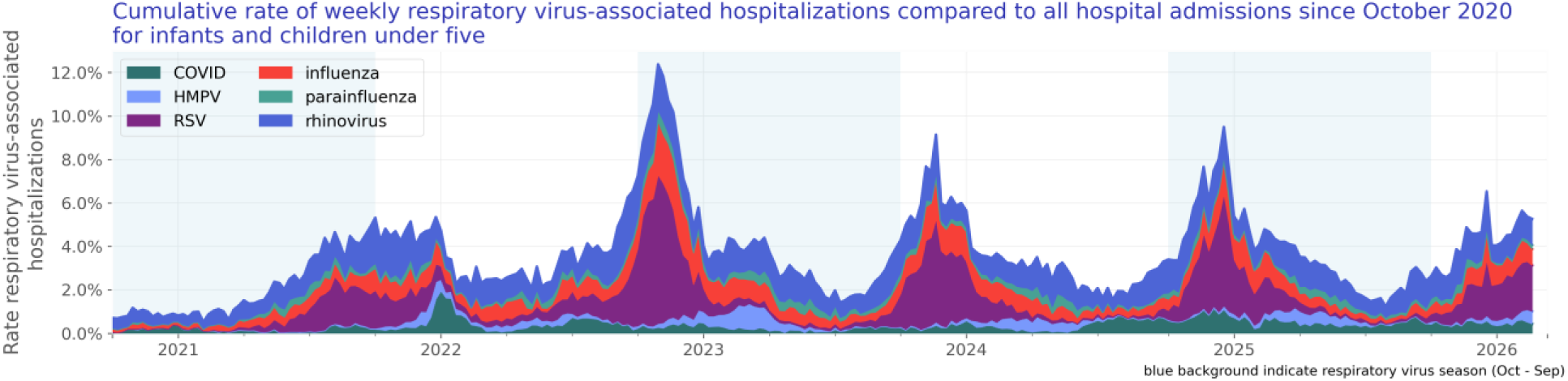
Cumulative rate of weekly respiratory virus-associated hospitalizations compared to all hospital admissions since October 2020 for infants and children under five.

#### Test positivity rate over time

We included 9,028,417 lab results with known results of infants and children under age five. Of those tests, 1,051,827 lab results were positive. The test positivity rate for infants and children under age five is shown in Figure 30.

**Figure 30:**
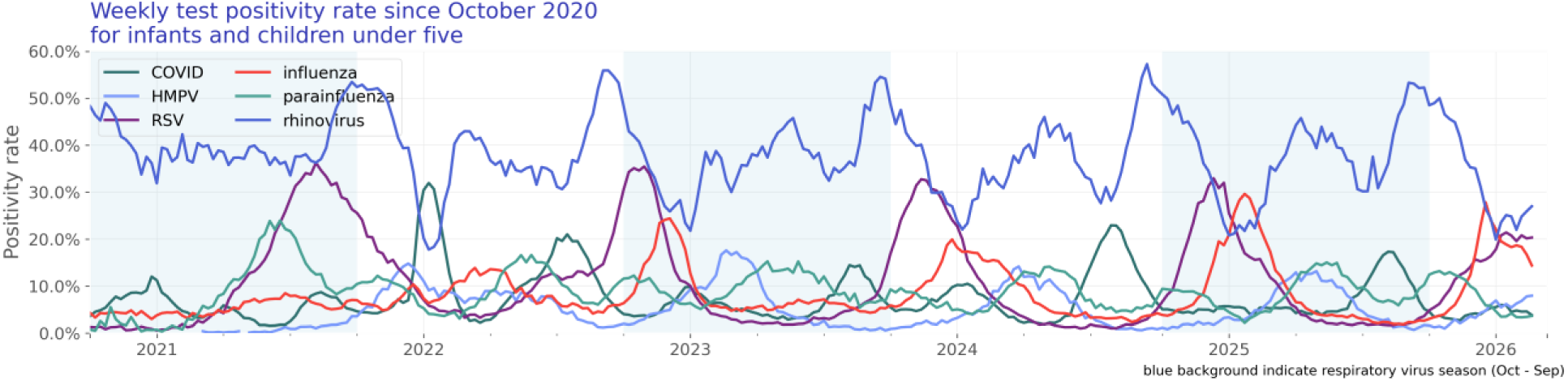
Weekly test positivity rate since October 2020 for infants and children under five.

### Older adults (age 65 and over)

Our older adults study population consists of 548,043 hospitalizations of 489,436 unique patients (Table 9).

**Table 9:** Older adults patient characteristics by season.

|  | 2020/2021 | 2021/2022 | 2022/2023 | 2023/2024 | 2024/2025 | 2025/2026 | Overall |
| --- | --- | --- | --- | --- | --- | --- | --- |
| N | 97,516<br>(100.00%) | 111,703<br>(100.00%) | 88,886<br>(100.00%) | 88,637<br>(100.00%) | 72,147<br>(100.00%) | 30,547<br>(100.00%) | 489,436<br>(100.00%) |
| Age Group |  |  |  |  |  |  |  |
| 65 - 74 years | 42,696<br>(43.78%) | 44,029<br>(39.42%) | 30,149<br>(33.92%) | 30,146<br>(34.01%) | 25,534<br>(35.39%) | 10,663<br>(34.91%) | 183,217<br>(37.43%) |
| 75 - 84 years | 35,349<br>(36.25%) | 41,738<br>(37.37%) | 34,678<br>(39.01%) | 35,065<br>(39.56%) | 28,563<br>(39.59%) | 11,895<br>(38.94%) | 187,288<br>(38.27%) |
| 85+ years | 19,471<br>(19.97%) | 25,936<br>(23.22%) | 24,059<br>(27.07%) | 23,426<br>(26.43%) | 18,050<br>(25.02%) | 7,989<br>(26.15%) | 118,931<br>(24.30%) |
| Sex |  |  |  |  |  |  |  |
| Female | 48,058<br>(49.28%) | 56,413<br>(50.50%) | 47,036<br>(52.92%) | 46,783<br>(52.78%) | 39,048<br>(54.12%) | 16,756<br>(54.85%) | 254,094<br>(51.92%) |
| Male | 49,048<br>(50.30%) | 54,863<br>(49.12%) | 41,491<br>(46.68%) | 41,483<br>(46.80%) | 32,894<br>(45.59%) | 13,577<br>(44.45%) | 233,356<br>(47.68%) |
| Unknown | 410 (0.42%) | 427 (0.38%) | 359 (0.40%) | 371 (0.42%) | 205 (0.28%) | 214 (0.70%) | 1,986<br>(0.41%) |
| Race |  |  |  |  |  |  |  |
| White | 71,543<br>(73.37%) | 84,687<br>(75.81%) | 66,849<br>(75.21%) | 65,390<br>(73.77%) | 53,955<br>(74.78%) | 22,637<br>(74.11%) | 365,061<br>(74.59%) |
| Black or African American | 10,546<br>(10.81%) | 11,330<br>(10.14%) | 8,674 (9.76%) | 9,506<br>(10.72%) | 7,240<br>(10.04%) | 3,377<br>(11.06%) | 50,673<br>(10.35%) |
| Asian | 1,661 (1.70%) | 1,823 (1.63%) | 1,938 (2.18%) | 2,022 (2.28%) | 1,760 (2.44%) | 816 (2.67%) | 10,020<br>(2.05%) |
| Native Hawaiian or Other Pacific Islander | 150 (0.15%) | 190 (0.17%) | 160 (0.18%) | 155 (0.17%) | 148 (0.21%) | 88 (0.29%) | 891<br>(0.18%) |
| American Indian or Alaska Native | 257 (0.26%) | 307 (0.27%) | 291 (0.33%) | 287 (0.32%) | 249 (0.35%) | 93 (0.30%) | 1,484<br>(0.30%) |
| Other Race | 3,208 (3.29%) | 2,930 (2.62%) | 2,821 (3.17%) | 3,127 (3.53%) | 2,881 (3.99%) | 1,166 (3.82%) | 16,133<br>(3.30%) |
| Unknown | 10,151<br>(10.41%) | 10,436<br>(9.34%) | 8,153 (9.17%) | 8,150 (9.19%) | 5,914 (8.20%) | 2,370 (7.76%) | 45,174<br>(9.23%) |
| Ethnicity |  |  |  |  |  |  |  |
| Hispanic or Latino | 4,855 (4.98%) | 4,231 (3.79%) | 3,979 (4.48%) | 4,682 (5.28%) | 4,383 (6.08%) | 1,769 (5.79%) | 23,899<br>(4.88%) |
| Not Hispanic or Latino | 81,044<br>(83.11%) | 96,223<br>(86.14%) | 76,410<br>(85.96%) | 75,774<br>(85.49%) | 62,204<br>(86.22%) | 26,741<br>(87.54%) | 418,396<br>(85.49%) |
| Unknown | 11,617<br>(11.91%) | 11,249<br>(10.07%) | 8,497 (9.56%) | 8,181 (9.23%) | 5,560 (7.71%) | 2,037 (6.67%) | 47,141<br>(9.63%) |
| Virus |  |  |  |  |  |  |  |
| COVID | 91,860<br>(94.20%) | 99,287<br>(88.88%) | 59,737<br>(67.21%) | 53,897<br>(60.81%) | 25,730<br>(35.66%) | 6,884<br>(22.54%) | 337,395<br>(68.94%) |
| HMPV | 31 (0.03%) | 1,310 (1.17%) | 2,505 (2.82%) | 2,323 (2.62%) | 3,207 (4.45%) | 1,393 (4.56%) | 10,769<br>(2.20%) |
| RSV | 558 (0.57%) | 1,667 (1.49%) | 3,411 (3.84%) | 5,763 (6.50%) | 6,326 (8.77%) | 3,148<br>(10.31%) | 20,873<br>(4.26%) |
| influenza | 2,682 (2.75%) | 5,792 (5.19%) | 16,435<br>(18.49%) | 18,779<br>(21.19%) | 26,480<br>(36.70%) | 13,883<br>(45.45%) | 84,051<br>(17.17%) |
| parainfluenza | 555 (0.57%) | 1,061 (0.95%) | 2,238 (2.52%) | 2,678 (3.02%) | 3,170 (4.39%) | 1,045 (3.42%) | 10,747<br>(2.20%) |
| rhinovirus | 1,830 (1.88%) | 2,586 (2.32%) | 4,560 (5.13%) | 5,197 (5.86%) | 7,234<br>(10.03%) | 4,194<br>(13.73%) | 25,601<br>(5.23%) |
| Comorbidity |  |  |  |  |  |  |  |
| Asthma | 7,612 (7.81%) | 9,881 (8.85%) | 9,495<br>(10.68%) | 10,337<br>(11.66%) | 9,653<br>(13.38%) | 4,334<br>(14.19%) | 51,312<br>(10.48%) |
| Chronic Lung Disease | 7,154 (7.34%) | 8,736 (7.82%) | 7,944 (8.94%) | 8,284 (9.35%) | 8,195<br>(11.36%) | 3,513<br>(11.50%) | 43,826<br>(8.95%) |
| Chronic Obstructive Pulmonary Disease | 17,015<br>(17.45%) | 23,921<br>(21.41%) | 21,145<br>(23.79%) | 22,825<br>(25.75%) | 20,294<br>(28.13%) | 8,590<br>(28.12%) | 113,790<br>(23.25%) |

#### Hospitalization rate over time

The rate of respiratory virus-associated hospitalizations compared to all hospitalizations for adults 65 and over is shown in ‘Figure 31. Figure 32 shows the same data stacked to represent the combined impact of the viruses.

**Figure 31:**
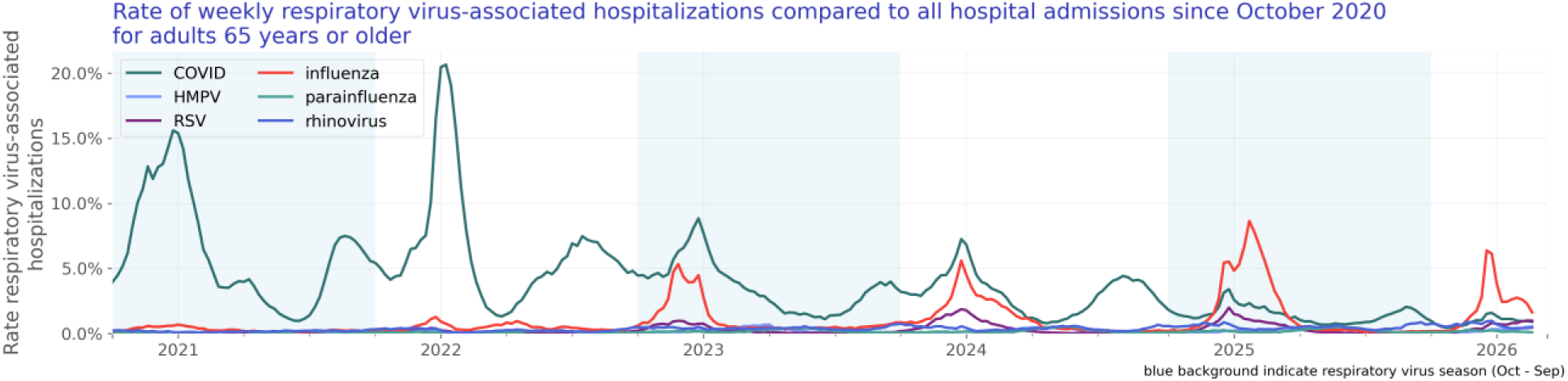
Rate of weekly respiratory virus-associated hospitalizations compared to all hospital admissions since October 2020 for adults 65 years or older.

**Figure 32:**
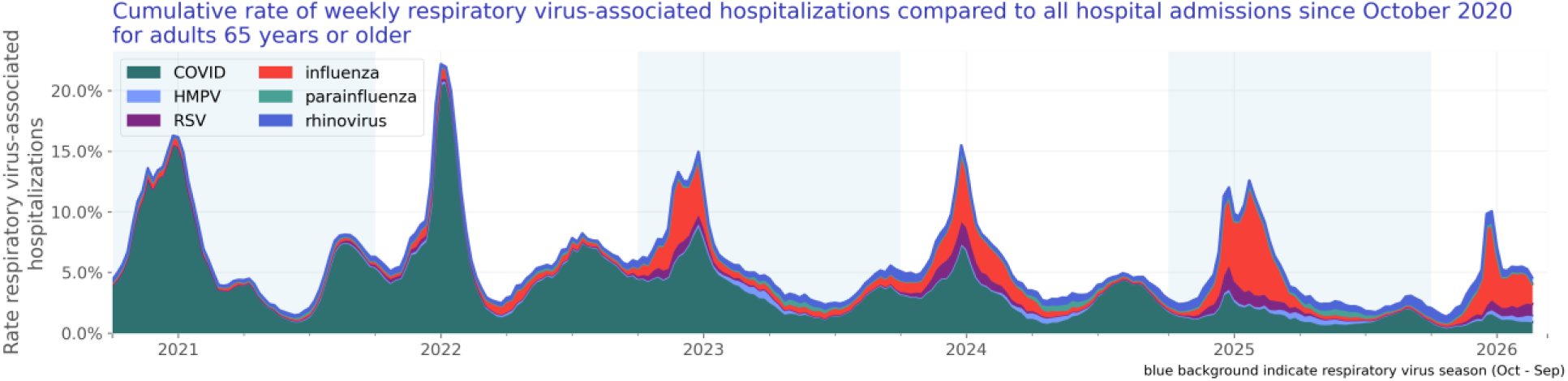
Cumulative rate of weekly respiratory virus-associated hospitalizations compared to all hospital admissions since October 2020 for adults 65 years or older.

#### Test positivity rate over time

We included 23,138,076 lab results with known results of adults aged 65 and over. Of those tests, 1,598,988 lab results were positive. The test positivity rate for adults aged 65 and over is shown in Figure 33.

**Figure 33:**
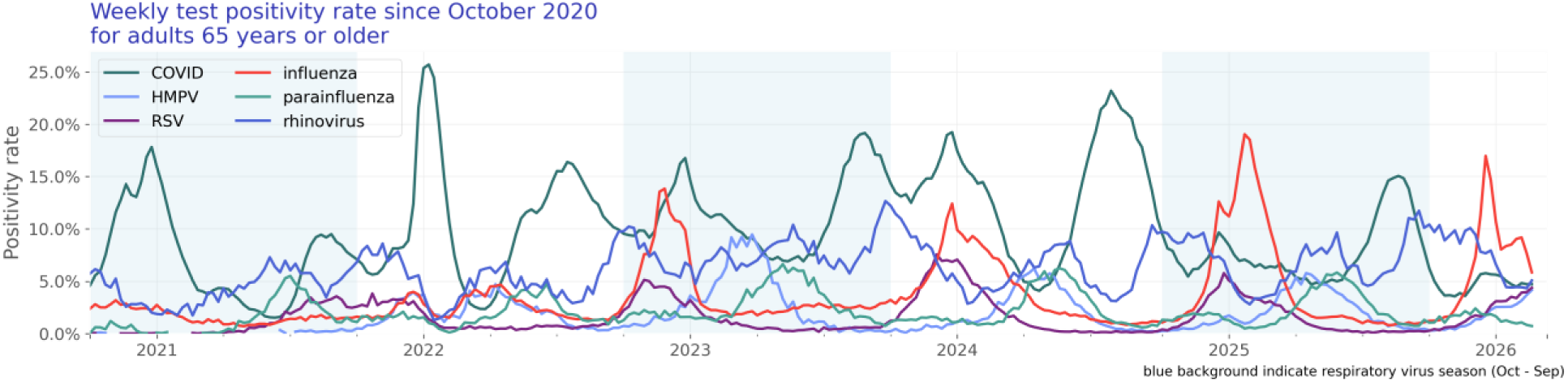
Weekly test positivity rate since October 2020 for adults 65 years or older.

## Discussion and conclusion

At the end of February 2026, respiratory virus-associated hospitalizations accounted for 3.6% of all hospitalizations, a slight decrease from the end of January 2026 (4.3%, - 15.3%). Influenza-associated hospitalizations decreased from 2.1% to 1.4% of all hospitalizations (-34.2%) and COVID-associated hospitalizations decreased slightly from 0.8% to 0.6% of all hospitalizations (-16.0%). RSV, rhinovirus, HMPV-, and parainfluenza-associated hospitalizations all remained relatively low and stable across the month.

Influenza, COVID, and parainfluenza test positivity decreased slightly (-14.3%, -13.5%, - 15.6%, respectively), while rhinovirus, RSV, and HMPV test positivity increased slightly (+11.8%, +9.2%, +64.1%, respectively).

The overall findings suggest a slight seasonal decline. Although influenza-associated hospitalizations declined, influenza still accounted for the largest proportion of respiratory virus–associated hospitalizations.

In the population aged 0–4 years, respiratory virus-associated hospitalizations increased slightly throughout February and are now at a rate of 4.3% of all hospitalizations in this age group (+11.8%). This increase was primarily driven by HMPV, with HMPV-associated hospitalizations more than doubling throughout February (+111.7%). RSV-, rhinovirus-, influenza-, COVID- and parainfluenza-associated hospitalizations remained stable (+7.6%, +5.4%, 7.5%, +17.9%, +91.6%). RSV-associated hospitalizations accounted for 2.1% of all hospitalizations by the end of the month, representing the largest share of respiratory-virus associated hospitalizations in this age group.

In this pediatric population, rhinovirus and RSV test positivity remained stable (+8.8% and -2.1%, respectively), influenza, COIVD, and parainfluenza test positivity decreased slightly (-21.4%, -15.2%, -13.1%, respectively), and HMPV test positivity increased (+46.8%). Together, these patterns suggest that HMPV drove the increase in respiratory virus activity among children aged 0–4 years, while activity for other viruses remained relatively stable.

Among adults aged 65 years and older, respiratory virus-associated hospitalizations accounted for 4.4% of all hospitalizations, a slight decrease from the end of January 2026 (5.3%, -17.0%). Influenza- and COVID-associated hospitalizations declined in February (-38.3% and -18.7%, respectively), while RSV-associated hospitalizations increased (+22.6%). HMPV, rhinovirus, and parainfluenza remained low and stable across the month.

In this older adult population, influenza and parainfluenza test positivity decreased (-31.2% and -32.6%, respectively), while rhinovirus, RSV, and HMPV test positivity increased (+15.1%, +24.4%, +61.5%). COVID test positivity remained stable across the month (+0.0%). Overall, these findings indicate a slight decrease in respiratory virus-associated hospitalizations among older adults in February, driven primarily by declines in influenza- and COVID-associated hospitalizations.

Several limitations of our analysis should be noted. All data are preliminary and may change as additional data are obtained. These findings are consistent with data accessed March 11, 2026. These are raw counts and post-stratification methods have not been conducted. This analysis does not include patients hospitalized with a respiratory virus who were not tested or were tested later in their medical care (when laboratory tests results would have returned a negative result). In addition, cohorts with small counts may be suppressed during the de-identification process leading to the appearance of zero patients for a given time period. Moreover, the unknowns in this report either indicate the value was not included in the individual’s electronic health record or that it was excluded from the data to protect an individual’s identity as a part of Truveta’s commitment to privacy (Truveta, 2024).

We will continue to monitor respiratory virus-associated hospitalization overall and for the at-risk populations.

## Data Availability

The data used in this study are available to all Truveta subscribers and may be accessed at studio.truveta.com.

## Notes

### Competing Interest Statement

All authors are employees of Truveta, Inc.

### Funding Statement

This study did not receive any funding.

### Author Declarations

Normalized electronic health record data are de-identified by expert determination under the HIPAA Privacy Rule before being made available to researchers. In accordance with 45 C.F.R. Para. 46.101 Protection of Human Subjects, our study did not require Institutional Review Board approval because it used only deidentified medical records. All data used in this study are publicly available to Truveta subscribers and may be accessed at studio.truveta.com.

### Summary of Updates

Data are updated through February 2026.

